# Pharmacokinetics, pharmacodynamics, efficacy and drug resistance selection of injectable long-acting lenacapavir pre-exposure prophylaxis (PrEP) against HIV

**DOI:** 10.1101/2025.08.26.25334527

**Authors:** Hee-yeong Kim, Antonia Liebenberg, Lanxin Zhang, Max von Kleist

**Affiliations:** Project group 5 “Systems Medicine of Infectious Disease”, Robert Koch Institute, Berlin, Germany; Mathematics for Data Science, Dep. of Mathematics and Computer Science, Freie Universität Berlin, Germany

## Abstract

Oral pre-exposure prophylaxis (PrEP) can substantially reduce HIV infection risk when taken as prescribed. However, many individuals struggle adhering to the daily regimen. Twice-yearly injections of the novel HIV capsid inhibitor lenacapavir (LEN) demonstrated potential in recent PrEP-trials. However, clinical trials may not enable to accurately estimate efficacy or protective concentration benchmarks. Moreover, while LEN can persist for more than a year, stopping PrEP may facilitate de novo drug resistance emergence.

We developed an integrated PK-PD model of LEN, incorporating PK-variability to quantify prophylactic efficacy against wild-type virus and transmitted drug resistance and to estimate the probability of drug resistance emergence when LEN-PrEP is stopped.

We estimated a 95% preventive and fully preventive plasma concentration of 4.7ng/mL and >5ng/mL, respectively. The latter was reached within 23hours after the first 927mg LEN SC injection and maintained up to 50.5weeks after the last dose in an ‘average’ individual. Considering PK-variability, concentrations of >5ng/mL were not consistently maintained at all times for lower concentrations, but were surpassed at steady-state. Full protection was achieved at 21, 59, 1108, 142, 538, 107, 1142ng/mL for viruses carrying mutations Q67H, N74D, Q67H+N74D, Q67H+T107N, M66I+T107A, Q67H+K70R, Q67H+K70R+T107N, respectively, and mutant selection windows for N74D, all double mutants and Q67H+K70R+T107N overlapped with LEN SC steady-state concentrations. In an ‘average’ individual, wild-type infection with subsequent de novo resistance emergence may occur within a period of ≈206, 170, 138, 160, 106, 191, 235days for Q67H, N74D, Q67H+N74D, Q67H+T107N, M66I+T107A, Q67H+K70R, Q67H+K70R+T107N after stopping LEN-injections, calling for strategies to manage LEN-PrEP discontinuation.

## Introduction

HIV remains a relevant public health treat, with globally 1.3 million new infections, 39.9 million people living with HIV (PWHIV) and 630,000 HIV-related deaths in 2023 [1]. While highly active antiretroviral treatment (HAART) can suppress virus replication and prevent acquired immunodeficiency syndrome (AIDS) [2], HIV persists in latently infected cells and may immediately rebound when treatment is stopped [3]. Moreover, there is no highly effective HIV vaccine to date. While vaccine options are missing, many antivirals can also be used as pre-exposure prophylaxis (PrEP) to prevent HIV infection. HIV PrEP with once-daily oral tenofovir disoproxil fumerate and emtric-itabine (TDF/FTC) is highly cost-effective [4], widely available and highly efficient in preventing HIV infection, when taken regularly [5]. However, adhering to the daily oral regimen poses challenges to some individuals, and in particular for heterosexual cis-women, where most HIV infections occur globally [6]. For example, in recent PrEP clinical trials [7], as little as 16% of study participants adhered to oral TDF/FTC by week 52.

Long-acting (LA-)PrEP formulations might offer a solution to individuals struggling to adhere to a once-daily regimen. Bi-monthly injections with carbotegravir (CAB) [8, 9] proved efficient in preventing HIV infection in both MSM and cis-gender women and are available in many high-income countries. Trials with islatravir implants for PrEP have been put on halt, due to side effects [10], while, more recently, twice-yearly injections with long-acting lenacapavir (LEN) successfully completed Phase III clinical testing, indicating > 90% prophylactic efficacy [7, 11]. However, it is challenging to precisely quantify the extent of LEN prophylactic efficacy, because of statistical limitations of clinical PrEP studies, as well as limitations in their design [12, 13]. Moreover, the risk of drug resistance emergence is difficult to systematically evaluate in clinical trials. While LA-PrEP offers the advantage of infrequent dosing, the long persistence of antiviral drugs may pose considerable risk to drug resistance selection, particularly when LA-PrEP is stopped due to side effects, lack of financial means, insurance coverage or unwillingness to continue PrEP. For example, CAB and LEN have plasma half-lives of ≈ 47 and 56-84 days, respectively [14], implying that drug concentrations insufficient to protect from infection, but sufficient to select drug resistance, may persist up to 1 year after stopping LA-PrEP. The case of an individual who had to stop LA-CAB, because he fell out of social insurance, acquired HIV infection and subsequently developed de novo drug resistance should be interpreted as a warning in this context [15].

LEN is a first-in-class inhibitor of HIV capsid function [16, 17] and used as a salvage therapy in heavily treatment-experienced adults with multi-drug resistant HIV, because there are no known cross-resistances to other antiretrovirals [18] . However, the use of LEN in salvage therapy makes evaluations regarding drug resistance selection and propagation particularly relevant.

Oral and subcutaneous (SC) formulations of LEN were in clinical development, with two recommended initiation regimens available for treatment and prophylaxis. These involved an oral lead-in phase with 600mg LEN as tablets, followed by SC injections of 927mg every six months. In addition, a new intramuscular (IM) formulation is being investigated in a Phase I clinical trial for PrEP. This once-yearly regimen includes a 5000mg IM dose of LEN and is being evaluated in two formulations (containing 5% w/w and 10% w/w ethanol) [19]. LEN has a relatively low barrier to drug resistance and *in vitro* resistance selection experiments have identified seven major LEN-associated mutations (L56I, M66I, Q67H, K70N, N74D, N74S, T107N and their combinations) affecting LEN binding to the capsid protein [20, 21]. Of those mutations, N74D was detected in 2/2 participants who got infected during the PURPOSE 2 PrEP-study [11].

Currently, no mathematical model has been published to quantify prophylactic efficacy of LEN for different administration schemes, or for assessing the risk of drug resistance emergence in the context of LA-LEN PrEP. In order to fill this knowledge gap, we developed an integrated PK/PD model for LEN derived from a published pharmacokinetic model [22] and was informed by publicly available clinical data from oral administration, long-acting subcutaneous (SC) and intramuscular (IM) formulations. An HIV-1 viral dynamics model was used to estimate the antiviral potency (*IC*_50_) of LEN against wild-type (WT) virus. We extended the viral dynamics model to account for the emergence and dynamics of drug-resistant variants utilizing *in vitro* phenotypic data of clinical isolates [20]. We then evaluated the efficacy of long-acting LEN across a range of dosing regimen against wild-type virus and transmitted drug resistance. Finally, we assessed the risk of de novo drug resistance emergence after wild-type infection, in case when LA-LEN PrEP is stopped. Overall, our study provides a quantitative framework to assess the benefits and potential risks of LEN with regards to PrEP and drug resistance selection.

## Methods

### Clinical PK data

We considered all relevant, publicly available pharmacokinetic data, as detailed in Supplementary Table S1. In total, this dataset encompassed 15 different dosing schemes and three administration routes (oral, subcutaneous and intramuscular) from which we extracted mean and median pharmacokinetic data (Engauge Digitizer).

### LEN PK modeling

#### LEN Compartment Model

Mean or median concentration–time data were fitted to a one-compartment pharmacokinetic (PK) model that can receive input from either oral administration, or one of the two parenteral routes following subcutaneous or intramuscular injection of LEN, Fig. 1A, akin to [22] . In brief, LEN in blood plasma is eliminated with rate constant *k*_*e*_. The oral route follows first-order absorption kinetics with rate *k*_*a*_. Subcutaneous (SC, abdominal) or intramuscular (IM, gluteal) injection involves parallel release processes: a fraction of the injected dose (*Frac* · dose_inject_) is released by a direct mechanism (with rate *k*_*direct*_) from the soluble fraction of the formulation. The remaining part is released by an indirect mechanism (with rate *k*_*indirect*_) through the formation of a solid depot at the injection site. As a result, part of the dose enters systemic circulation rapidly, while the solid depot dissolves gradually over time, ensuring steady drug levels over a prolonged period. The directly absorbed fraction at the first dosing time (*t* = 0) was modeled by *PI*_*dir*_(0) = *Frac* · dose_inject_. To account for the delay in systemic availability (lag time), the indirect release pathway was modeled using *n* transit compartments with identical first-order transit rates, corresponding to an Erlang distribution [22, 23] with initial state *PI*_*ind*_(0) = (1 − *Frac*) · dose_inject_. All parameter estimates are listed in Table 1. The corresponding ODEs are given below.

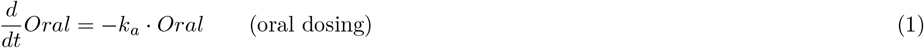

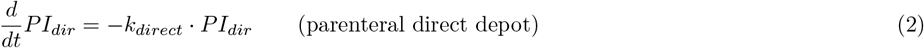

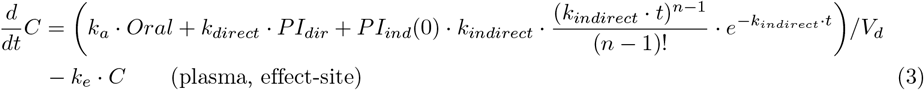

**Table 1.** LEN mean PK parameter estimates. We found that a power-law function *f* (dose) = 508.16 · dose^0.61^ with input dose in mg can be used to derive dose specific *V*_*d*_ values. More details see section *Methods* [22].

| Parameter | ID: Model estimates | Unit | Description |
| --- | --- | --- | --- |
| $k_a$ | Study 2: 1.08 | 1/h | Oral absorption rate constant.<br>Numerically obtained using Eq. (4). |
| $k_{direct}$ | Study 1: 0.1<br>Study 3: 0.063<br>Study 4: 0.039<br>Study 5: $2.6 \times 10^{-4}$ | 1/h | Direct absorption rate constant.<br>Parameters obtained by optimization. |
| $k_{indirect}$ | Study 1: $6.5 \times 10^{-4}$<br>Study 3: 0.0052<br>Study 4: $6.3 \times 10^{-4}$<br>Study 5: $4.1 \times 10^{-4}$ | 1/h | Indirect absorption rate constant.<br>Parameters obtained by optimization. |
| $Frac$ | Study 1: 12<br>Study 3: 16<br>Study 4: 1.5<br>Study 5: 12.8 (F1), 43.6 (F2) | % | Direct fraction of parenteral injections.<br>Parameters obtained by optimization. |
| $n$ | Study 1: 1<br>Study 3: 3<br>Study 4: 2<br>Study 5: 2 | - | Number of transit compartments. |
| $V_d$<br>$V_d(300)$<br>$V_d(900)$<br>$V_d(1,800)$ | 1618<br>13304<br>35374<br>46516 | L | Volume of distribution for parenteral<br>and oral dosing. Parameters<br>obtained by optimization. |
| $k_e$ | 0.0026 | 1/h | Global elimination rate constant.<br>Graphically determined from<br>log-transformed oral PK data. |

**Fig 1.**
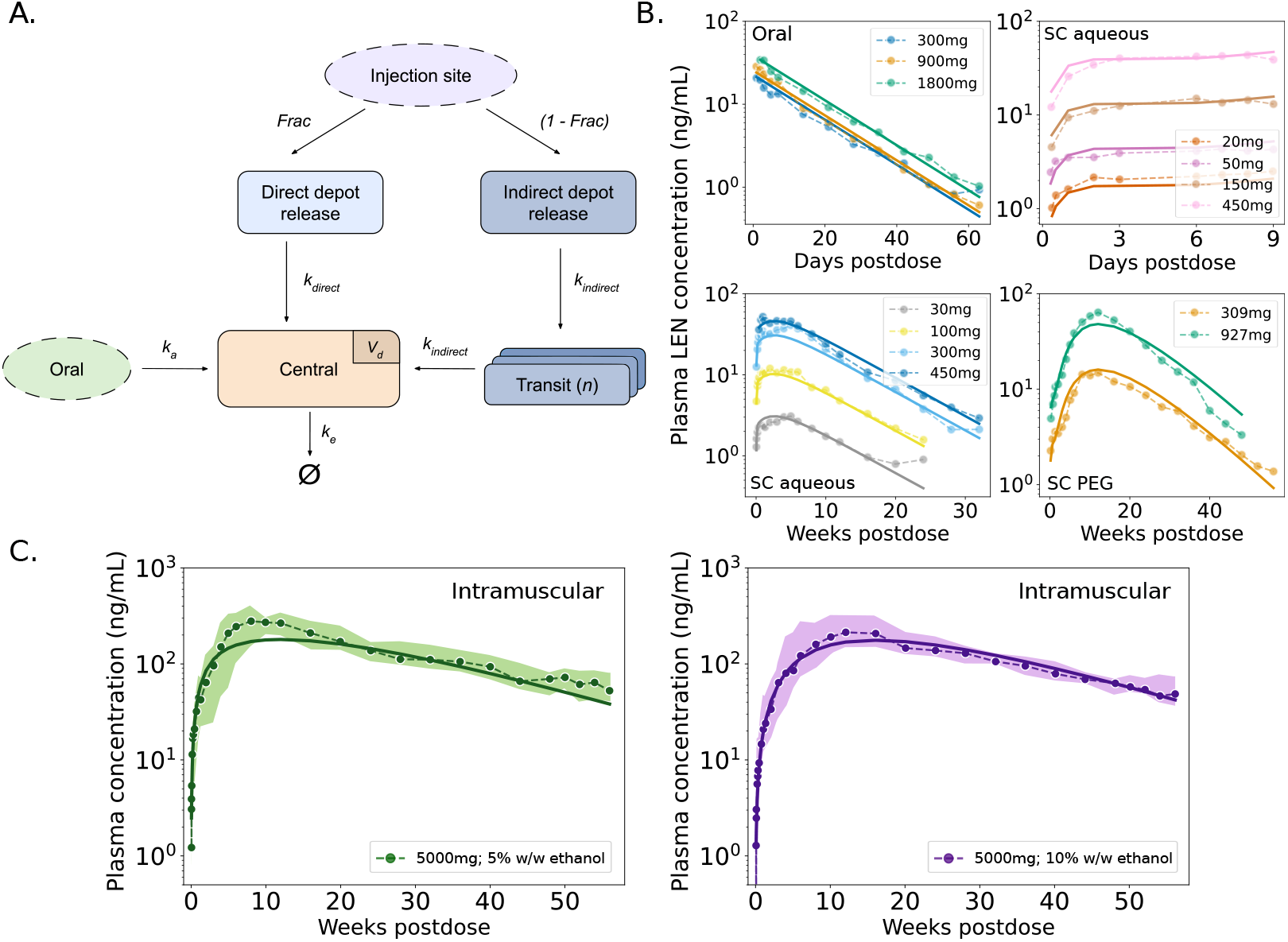
Pharmacokinetics of LEN. A. Schematic representation of the developed PK-model for LEN. The model includes first-order absorption for oral dosing (tablets) and dual-release kinetics (direct and indirect depot release) for parenteral formulations, including subcutaneous (SC) and intramuscular (IM) injections. B. Model fitting results for LEN plasma concentrations after oral administration and three distinct SC formulations. Dots represent average concentrations observed in the respective clinical studies, and model predictions are shown as continuous lines based on best-fit parameters. C. Model-predicted PK profile for long-acting, next-generation intramuscular (IM) injection with LEN, formulated with 5% and 10% w/w ethanol. Dots show clinical median concentrations, and shaded areas represent the interquartile range (IQR; from the first to the third quartile from [19]). Clinical data used for the fitting are summarized in Supplementary Table S1. Model performance was evaluated based on the reported PK properties listed in Supplementary Table S2.

#### LEN Oral PK

The determination of the absorption rate constant, *k*_*a*_, is based on the time to maximum concentration (*T*_*max*_), assuming a one-compartment model with linear absorption and first-order elimination. Dose-dependent *T*_*max*_ values were obtained from the respective PK study, see Supplementary Table S2. The plot of the log-transformed PK data vs. time is linear, and the slope yields the elimination rate constant *k*_*e*_. The absorption rate constant *k*_*a*_ could then be numerically obtained from the following equation:

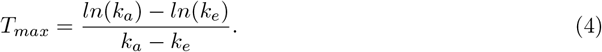

Plasma levels tended to increase with higher doses disproportionally with dose. Non-linear plasma levels were modeled with a dose-specific *V*_*d*_(dose) [22].

#### LEN Parenteral PK

LEN exhibits flip-flop PK [22], characterized by a slower absorption compared to the elimination rate (i.e. *k*_*direct*_ +*k*_*indirect*_ *< k*_*e*_). This makes absorption the rate-limiting factor of the terminal elimination phase. To maintain the kinetics, we fitted individual absorption parameters for the three SC and two IM parenteral formulations. This includes the fraction of drug directly entering systemic circulation (*Frac*) and the number of transit compartments (*n*) to capture the lag time, while we estimated one volume of distribution *V*_*d*_ across all parenteral formulations and fixed the elimination rate *k*_*e*_ to the value obtained from oral dosing. The SC-PK data show an approximately dose-proportional increase in exposure, whereas the available IM data were not sufficient to obtain a relationship. All parenteral formulations show distinct plasma pharmacokinetics (see Table 1). In each case, the indirect absorption pathway is much slower than the direct process, contributing to a prolonged half-life (shown in Supplementary Table S2).

#### Numerical simulation

For numerical simulation of LEN pharmacokinetics, we solved the system of ordinary differential equations (eqs. (1)-(3)) with corresponding initial conditions *z*_0_ = [dose_oral_, 0, 0] for oral administration and *z*_0_ = [0, *Frac* · dose_inject_, 0] for SC and IM administration using *scipy*.*integrate*.*solve ivp()*, version 1.16.0 and LSODA as numerical solver.

Multiple doses were modeled by adding the dose at each predefined dosing event time *τ*_*j*_ to the dosing compartment *D*, depending on the route of administration: 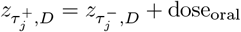 for oral dosing, and 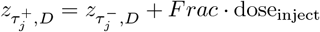 for SC/IM direct absorption. The remaining fraction of the SC/IM dose (1 − *Frac*) · dose_inject_ was added to the Erlang input function, describing the indirect absorption pathway, which was evaluated as a function of *t* − *τ*_*j*_ (time post-dosing). Here, 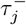 represents the time of the dosing event (before applying the dose) and 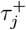 after applying the dose.

#### Parameter estimation

For oral dosing, we estimated a single dose-dependent parameter (volume of distribution *V*_*d*_). Based on our analysis, this parameter can also be obtained by the power-law function *f* (dose) = 508.16 · dose^0.61^ with input dose in mg. For SC and IM dosing, 4 model parameters were estimated, including a common *V*_*d*_ and fixed *k*_*e*_. Parameters *k*_*direct*_, *k*_*indirect*_ and *Frac* were optimized for a fixed number of transit compartments *n*. The value *n* was varied (*n* ∈ {1, …, 5}) and parameter optimization was repeated for each fixed *n*. All free parameters were optimized in a least-squares sense using *lmfit*.*minimize()* with basin-hopping global search (SLSQP as local optimizer), version 1.3.3. The model equations were numerically integrated using SciPy’s *solve ivp()* and LSODA. Final parameter estimates are depicted in Table 1.

#### PK variability

To account for inter-individual variability (IIV), we extracted data (Engauge Digitizer) from the Purpose 2 trial, which assessed 927 mg twice yearly SC injections and recorded concentration measurements across a random cohort of 10% of study participants at weeks 4, 8, 13, 26, 39 and 52 after the first LEN injection [11]. Ratios of minimum and maximum concentrations relative to the median were calculated for these six time points, and the geometric mean of these ratios was then determined across all time points. This yielded a lower PK variability limit of 0.15 times the median concentration and a factor of 4.0 times the median concentration as an upper concentration limit. To depict PK variability, we applied these ranges (0.15×median, 4.0×median) to simulated data (SC and IM injections) from our PK model. An application of these ranges to data from Phase I single SC dose 927 mg injections is shown in Supplementary Fig. S1 together with the original PK variability data from Purpose 2.

### LEN antiviral effects

Based on LEN’s main mechanism of action (MOA), we modeled LEN to interfere with the late phase of the HIV-1 replication cycle, similar to protease and maturation inhibitors [24]. I.e., in our model LEN would disrupt viral maturation, leading to the production of non-infectious viral particles, as shown in Fig. 2.

**Fig 2.**
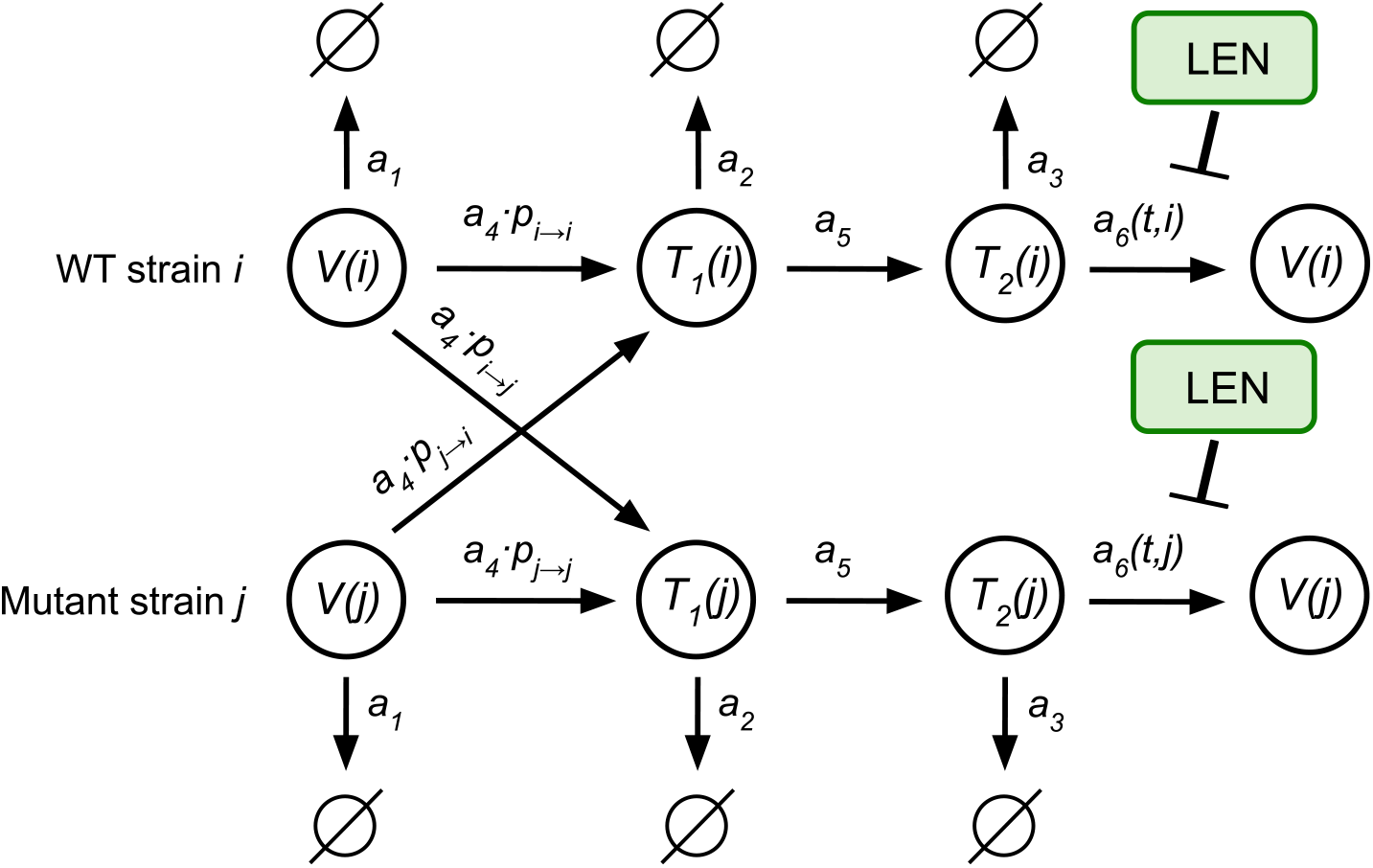
Simplified viral dynamics model incorporating the mechanisms of action of LEN and the emergence of mutated strains. Free infectious virus (*V*), early infected cells (*T*_1_) and late infected cells (*T*_2_) can be cleared with reaction rates 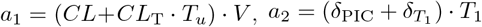and 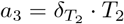 respectively. A susceptible T-cell (*T*_*u*_) can be successfully infected by an infectious virus at rate *a*_4_ = *β* · *T*_*u*_ · *V*, resulting in an early-infected T-cell, which may transition into a late-infected T-cell with rate *a*_5_ = *k* · *T*_1_. Mutations can emerge during reverse transcription (reaction *a*_4_). LEN inhibits the production of infectious virus (*V*), with reaction rate *a*_6_ = *s*(*i*) · (1 − *η*(*C*_*t*_, *i*)) *N*_T_ · *T*_2_. Above, *β* denotes the rate parameter associated with successful infection of *T*_*u*_ cells, while *CL*_T_ denotes the rate parameter for unsuccessful infection of HIV target cells. Free infectious virus gets cleared at rate parameter *CL*. Parameter *k*_*T*_ is related to the rate of successful integration of the viral genome, while 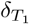and *δ*_*P IC*_ represents the rate parameters of clearance of early infected cells *T*_1_ and the intracellular destruction of the pre-integration complex (PIC) respectively. Parameters *N*_T_ and *δ*_*T* 2_ denote the rate at which infectious virus is produced by late infected T-cells and the clearance of late infected T-cells respectively. Parameters and derivations of this simplified viral dynamics model are given in Supplementary Table S3 and the Supplementary Text.

#### Estimation of antiviral potency

We used viral load (VL) data from a Phase Ib proof-of-concept study, that assessed the mean log_10_-transformed change in plasma HIV-1 RNA/mL following SC aqueous suspension administration (Study 3) of 20–450 mg LEN in individuals with untreated HIV-1 infection [17]. The decline in VL following LEN monotherapy was modeled by linking the PK model to an established HIV-1 viral dynamics (for details, see *Supplementary Text* and Supplementary Table S3 for parameters). To model direct drug effects *η*(*C*_*t*_, *FC*(*i*)), we utilized a standard Emax model:

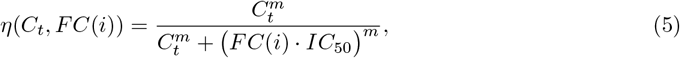

where *C*_*t*_ denotes the plasma LEN at time *t, IC*_50_ denotes the fifty percent inhibitory concentration against wild-type and the Hill coefficient was set to *m* = 2.1 [25]. *FC*(*i*) *>* 1 denotes the fold-change (*FC*) for resistant strains *i*. Mutation-specific values are presented in Supplementary Table S4. The viral dynamics parameters with *β*_*T*_ = 1.8 × 10^*−*12^ 1/day and *β*_*M*_ = 2 × 10^*−*14^ 1/day were adjusted to match the patients baseline VL of approximately 4.5 log_10_ HIV-1 RNA copies/mL in plasma. These parameters also align with the onset of the second-phase viral decay, both in the absence of LEN. After PK-PD coupling, the *IC*_50_ was estimated in a least-squares sense.

### Mutant selection window and probability of infection with resistant strains

During its elimination phase, LEN concentrations change very slowly. This means that LEN concentrations remain approximately constant at the timescale of infection establishment (or viral elimination). This time-scale separation allows to greatly simplify the computation of infection probabilities and mutant selection using the analytical solutions presented in the *Supplementary Text* and based on the strain-specific reproduction number.

The reproduction number *R*_0_(*i*) estimates the average number of infectious progeny produced by a (mutant) founder virus *i* during a single replication cycle and can be computed from a viral dynamics model. We simplified the viral dynamics model focusing on T-cells dynamics, as this is sufficient to estimate prophylactic efficacy [26], yielding the model depicted in Fig. 2 (derivations in *Supplementary Text*). We set the rate of mutations to 0 and defined a drug-independent coefficient:

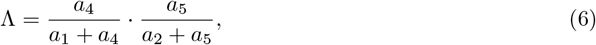

which captures the efficiency of target cell infection and integration up to the virus production-competent compartment. The reproduction number *R*_0_(*i*) for virus strain *i* is then given by:

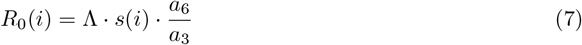

where *a*_6_(*i*)*/a*_3_ represents the average number of infectious virus being produced from a late infected T-cell (*T*_2_) before it is cleared. In line with classical results, *R*_0_ *<* 1 implies that the infection dies out, whereas *R*_0_ *>* 1 indicates that the virus replicates and may establish infection. Based on the utilized viral dynamics parameters from [27] (depicted in Supplementary Tab. S3), we computed *R*_0_(*WT*) ≈ 11.3 for the wild-type *WT* and in the absence of drugs.

In the presence of LEN concentrations *C*_*t*_, the instantaneous reproduction number of a viral strain *I* is then computed as

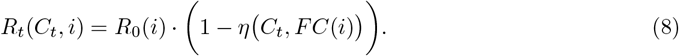

which allows to integrate PK/PD models (eqs. (1)-(3), eq. (5)). To model mutation-specific attributes, we used values for *s*(*i*) and *FC*(*i*) derived from *in vitro* phenotypic assays of clinical isolates, incorporating variability in fold-change and intrinsic fitness values (details in Supplementary Table S4).

When LEN concentrations remain approximately constant over the duration of the infection event, the infection probability, after a single virus reaches a replication-competent environment, can be calculated directly from the instantaneous reproduction number, akin to [28] (see *Supplementary Text* for further details):

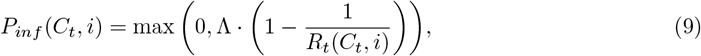

To calculate an average infection probability after homosexual virus exposure, the number of transmitted virions was drawn from a distribution relating donor virus loads to inoculum size in a replication-competent environment [29] and infection probabilities when *n >* 1 viruses were inoculated were computed as *P*_*inf,n>*1_(*C*_*t*_, *i*) = 1 − (1 − *P*_*inf*_ (*C*_*t*_, *i*))^*n*^, assuming statistical independence. We then calculated prophylactic efficacy as the relative HIV infection risk reduction relative to wild-type virus challenge in the absence of LEN.

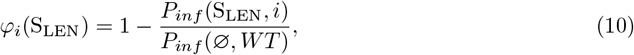

where *P*_*inf*_ (S_LEN_, *i*) denotes the probability of infection with mutant strain *i* in the presence of LEN and *P*_*inf*_ (∅, *WT*) denotes the probability of infection with WT virus in absence of prophylaxis. This allows to assess the impact of fitness deficits *s*(*i*) of mutant viruses *i*, as well as the impact of LEN and any drug resistance phenotype simultaneously. I.e., in the absence of LEN, mutants may not spread as efficiently if they have fitness deficits, whereas at increasing LEN concentrations selective pressure may favor mutants over the wild-type.

### Viral infection with de novo emergence of mutations

To study *de novo* emergence of mutations, we computed the infection probability after wild-type exposure, considering eventual *de novo* drug resistance dynamics. For this, we adapted the approach in [26] to our model in Fig. 2 to compute the probability of extinction in a multi-strain setting comprising wild-type (WT) and mutant strains. This yields the following set of ODEs,

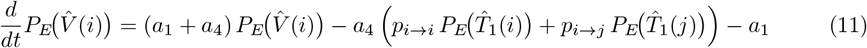

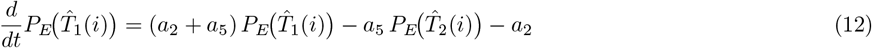

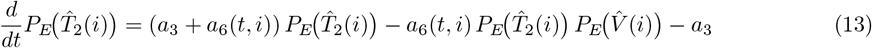

where 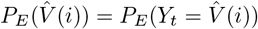 denotes the probability that a single mutant virion present at time *t* eventually goes extinct. The (time-varying) pharmacodynamic effect of LEN on mutant strain *i* is included in reaction *a*_6_(*t, i*) = *s*(*i*) · 1 − *η*(*C*_*t*_, *FC*(*i*)) · *a*_6_(*WT*, ∅) via the Emax model.

Parameter *p*_*i→j*_ = *µ*^*h*(*i,j*)^ · (1 − *µ*)^*N−h*(*i,j*)^ denotes the probability that any strain *i* mutates into another strain *j* where *h*(*i, j*) denotes the hamming distance between strain *i* and *j, N* denotes the total number of mutated positions and *µ* ≈ 2.16 · 10^*−*5^ represents the mutation probability per base [24].

After solving this system backwards in time using a standard ODE solver, the infection probability can be computed as

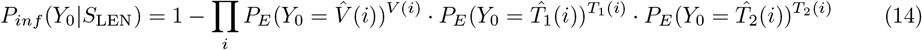

where *i* refers to a mutant strain and 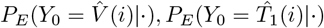and 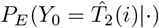 denote the probability of extinction upon exposure to a single free virus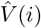 early infected cell 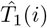 and late infected cell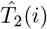, respectively. The exponents *V* (*i*), *T*_1_(*i*) and *T*_2_(*i*) represent the actual numbers that an individual was exposed to (assuming statistical independence). In our simulations regarding de novo drug resistance selection we have *V* (*WT*) *>* 0, while all other compartments were initiated with 0 particles and hence the equation above reduces to 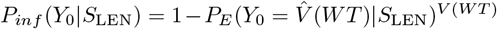 (more details, see *Supplementary Text*). Python codes for this method are linked via the GitHub repository stated in the Data and Code availability section.

## Results

### Pharmacokinetics and pharmacodynamics of LEN

We fitted a single pharmacokinetic (PK) model (Fig. 1A) to concentration-time data from 15 single dose regimens, including 3 administration routes (oral, subcutaneous (SC) and intramuscular (IM)), Supplementary Table S1. Predictions vs. average concentration-time profiles are shown in Fig. 1B-C indicating that the developed model appropriately captures average concentrations-time profiles across different doses and administration routes. The data and model highlight strong differences in drug uptake and distribution between oral and SC formulations, with a rapid uptake for oral regimen (*T*_max_ ≈ 4 − 5 hours) and a much longer time to peak concentrations for the different SC and IM formulations. Derived PK parameter values are summarized in Table 1, indicating slightly different parameter sets for the distinct formulations. Previously reported and model-predicted summary PK statistics (*C*_max_, *T*_max_ and *t*_1*/*2_) agreed very well as shown in Supplementary Table S2. Our modeling results highlighted that approximately 88% and 84% of the subcutaneously administered dose (SC aqueous suspensions) is released via an indirect absorption pathway (depot) in Studies 1 and 3, respectively. In Study 5 (IM formulation), the indirect release accounts for 88.8% for 5% w/w ethanol and 62.2% for 10% w/w ethanol, respectively. For Study 4 (SC PEG/water), the model predicted that 98.5% of the dose is released via a depot. Furthermore, LEN plasma concentrations may vary depending on the injection site and individual pharmacokinetic differences. Therefore, we extracted the possible range of injection-related variability from Phase III data and used it as baseline variability in our analysis (see Supplementary Fig. S1).

Using the parameterized PK model, we then fitted LEN’s antiviral potency by estimating viral decay kinetics from a single dose monotherapy study in HIV infected individuals (Supplementary Table S1; Study 3), accurately approximating viral load kinetics across four SC dosing regimen for which data was available, Fig. 3. Our parameter estimation resulted in an *IC*_50_ of 1.6ng/mL for direct target inhibition (IC_95_ = 6.5ng*/*mL), which is of the same order as the reported protein-adjusted *EC*_95_ = 3.87ng/ml [17]. Notably, the data (*n* = 6 per dosing regimen) indicated large inter-individual variability in viral decay, as also implicated by a larger average viral decay for 50 vs. 150 mg dosing. All considered doses led to an at least 10-fold reduction in plasma HIV-1 RNA (log_10_ copies/mL) through day 10 [17].

**Fig 3.**
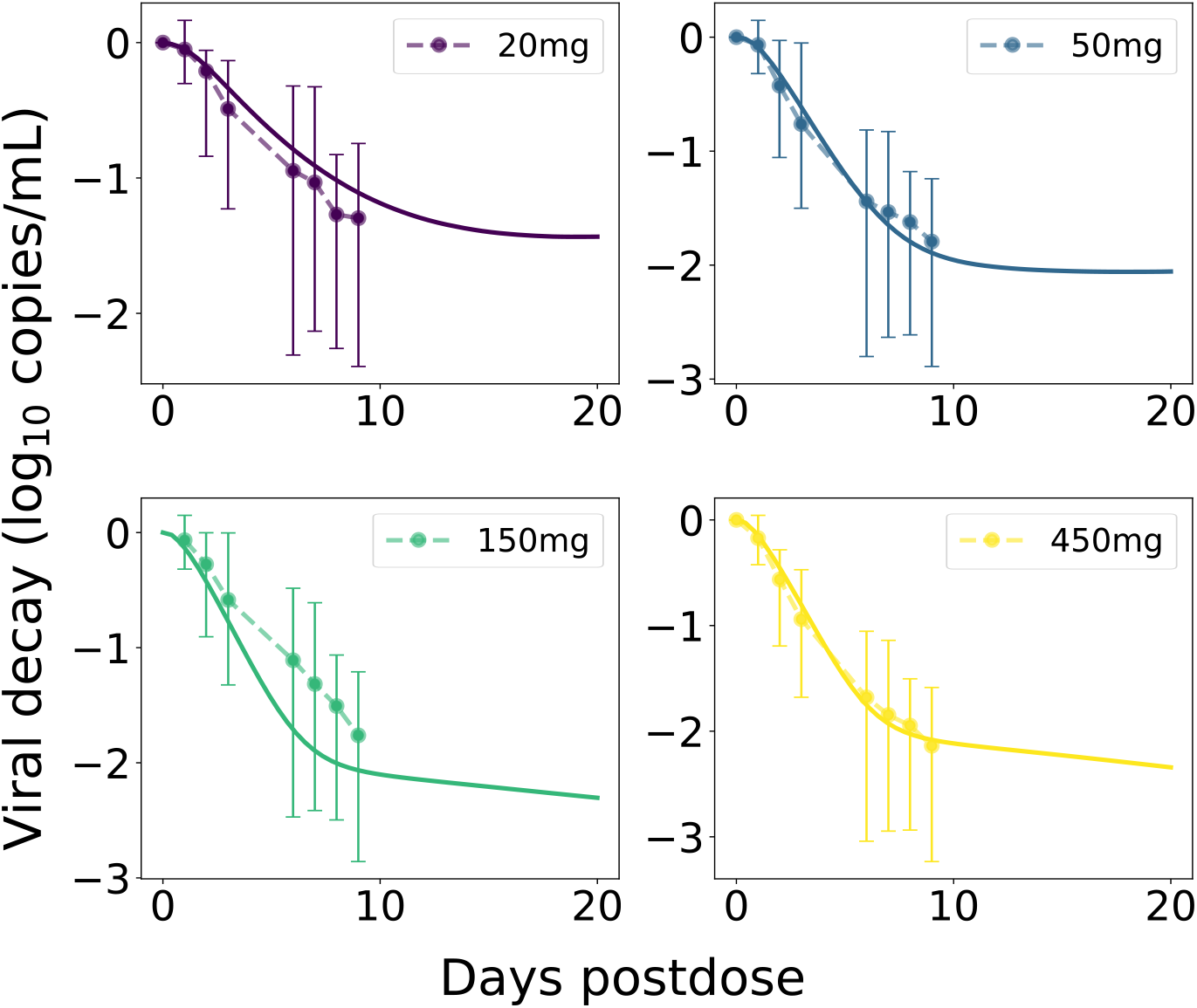
LEN viral decay in plasma with best-fit parameters. Average observed ‘change from baseline in HIV-1 RNA’ after single SC LEN injections (Study 3, aqueous suspension) are shown as dots with error bars representing minimum and maximum values. Model-predictions are shown as solid curves in the same color. The plasma VL (containing on average two viral RNAs [RNA/mL]) were calculated based on the total body virus, *V*_total_ = *V*_*I*_ + *V*_*NI*_, by assuming its distribution into plasma (*V*_plas_) and interstitial space (*V*_int_), with the volume of distribution determined as *K*_int:plas_ × *V*_int_ + *V*_plas_, where *K*_int:plas_ ≈ 50. For more details see [24].

### Analysis of mutant selection window

Next, we wanted to use the developed model to determine which concentrations of LEN may favor the selection of mutant strains. The selection of variants is influenced by fitness costs (i.e., selective disadvantage) and selection pressure through LEN, which may favor the resistant virus at high concentration ranges. The mutant selection window (MSW) is characterized by 1 *< R*_*t*_(*C*_*t*_, *WT*) ≤ *R*_*t*_(*C*_*t*_, *mut*) [30], i.e. the mutant virus replicates better than wild-type and it replicates to an extent that allows to sustain infection, compare Fig. 4A (hatched area). Both intrinsic mutant fitness (1 − *s*(*i*)) and fold-change *FC*(*i*) in drug susceptibility determine the size of the MSW.

**Fig 4.**
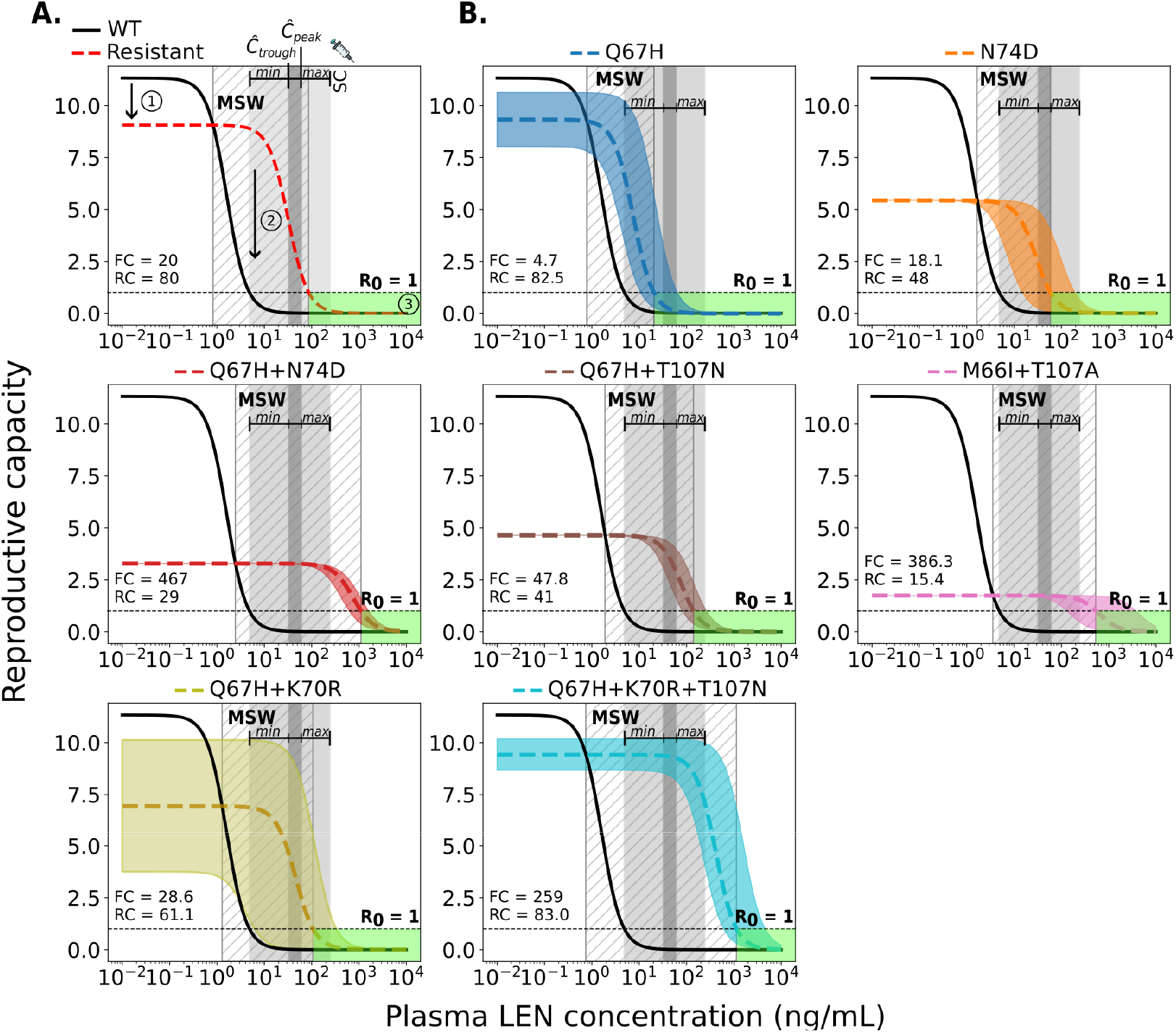
Relationship between plasma LEN concentrations after twice-yearly SC injections and viral reproductive capacity for wild-type and resistant strains. A. Example showing the impact of a resistant strain on viral fitness (red dashed line: 20% lower replication capacity) and drug susceptibility (solid black line: 20-fold higher *IC*_50_) compared to wild-type (WT). Key features: (1) reduced fitness of the mutant, (2) the mutant selection window (MSW, diagonal hatching) where the mutant outcompetes WT, and (3) the green area where reproductive capacity is ≤ 1 (infection cannot occur). B. MSW analysis of two single mutants, four double mutants, and one triple mutant. The horizontal dashed line at *R*_0_ = 1 marks the threshold between viral suppression (below 1) and sustained replication (above 1). The dark gray areas indicate the clinically relevant population-average steady-state concentration ranges (*C*_*trough*_-to-*C*_*peak*_ concentration ranges) achieved by twice-yearly SC LEN injections (PEG formulation). The light gray shaded areas represent ranges of inter-individual variability in drug concentrations informed by observations in the Purpose 2 trial (*min*-*max*). Each mutant (labeled at the top of each panel) is depicted as a colored dashed line, with uncertainty in fitness and/or fold-change represented by a corresponding shaded region.

We used *in vitro* phenotypic parameters [20] to calculate the MSW of SC LEN twice-yearly for single and double mutants, including a triple mutant, as shown in Fig. 4B. The single mutants Q67H and N74D show a relatively high to moderate fitness (1 − *s*(*i*) ≈ 82.5% and 48%), respectively. Q67H exhibits moderate resistance while maintaining high replication capacity (low selective disadvantage). According to our simulations, this mutation would be selected at LEN concentrations *>* 0.8ng/mL and would be sufficiently suppressed by concentrations exceeding 21ng/mL. At population-average *C*_through_ to *C*_peak_ concentration ranges (32-62ng/mL) following twice-yearly SC injections, Q67H would thus be sufficiently suppressed, whereas IIV concentration ranges (4.8-240ng/mL) may occasionally not fully suppress (and select) Q67H. On the other hand, peak-to-through concentration ranges fall within the MSW of mutant N74D (MSW_N74D_ ∈ [1.7, 59]). Regarding the single mutations, it is conceivable that Q67H may emerge under low drug pressure or pre-exist, with the additional N74D mutation appearing at higher concentrations or prolonged LEN exposure. The combination of both mutations leads to a highly resistant double mutant. If T107N or K70R were additionally selected, each would confer resistance while reducing replication capacity. However, this could favor the emergence of the triple mutant Q67H+K70R+T107N, which exhibits both high fitness and high resistance, as shown in Fig. 4B. The corresponding MSW analysis for once-yearly intramuscular LEN injections is shown in the Supplementary Fig. S2.

All evaluated double mutants and the triple mutation exhibited high levels of LEN resistance. The population-average steady-state (peak-to-through) concentration range following twice-yearly LEN injections (dark gray, Fig. 4) lies within the MSWs for Q67H+N74D, Q67H+T107N, M66I+T107A, Q67H+K70R and Q67H+K70R+T107N. The steady-state concentration range including inter-individual variability is fully contained within the MSWs of Q67H+N74D, M66I+T107A, and Q67H+K70R+T107N.

### Prophylactic efficacy of LEN against transmission of WT and drug resistant virus

Using the approach in eq. (9), we were able to estimate the relation between (static) LEN concentrations and prophylactic efficacy, Fig. 5. We observed a steep concentration-prophylaxis relationship, similar to that observed for protease- and maturation inhibitors in earlier works [28]. This response curve does not have a sigmoidal shape like the Emax equation typically used to model concentration-effect relationships, and therefore complicates the estimation of 95% and 99% preventive concentrations directly from *in vitro* inhibitory concentrations. When we calculated the drug concentration required to achieve 95% prophylactic efficacy (*EC*_95_) from the concentration-response curve depicted in Fig. 5, we obtained a value of *EC*_95_ = 4.7ng/mL against infection with wild-type (WT) virus. Moreover, according to our model, infection with WT virus would be fully averted at average steady-state (peak-to-through) LEN concentrations for twice-yearly SC.

**Fig 5.**
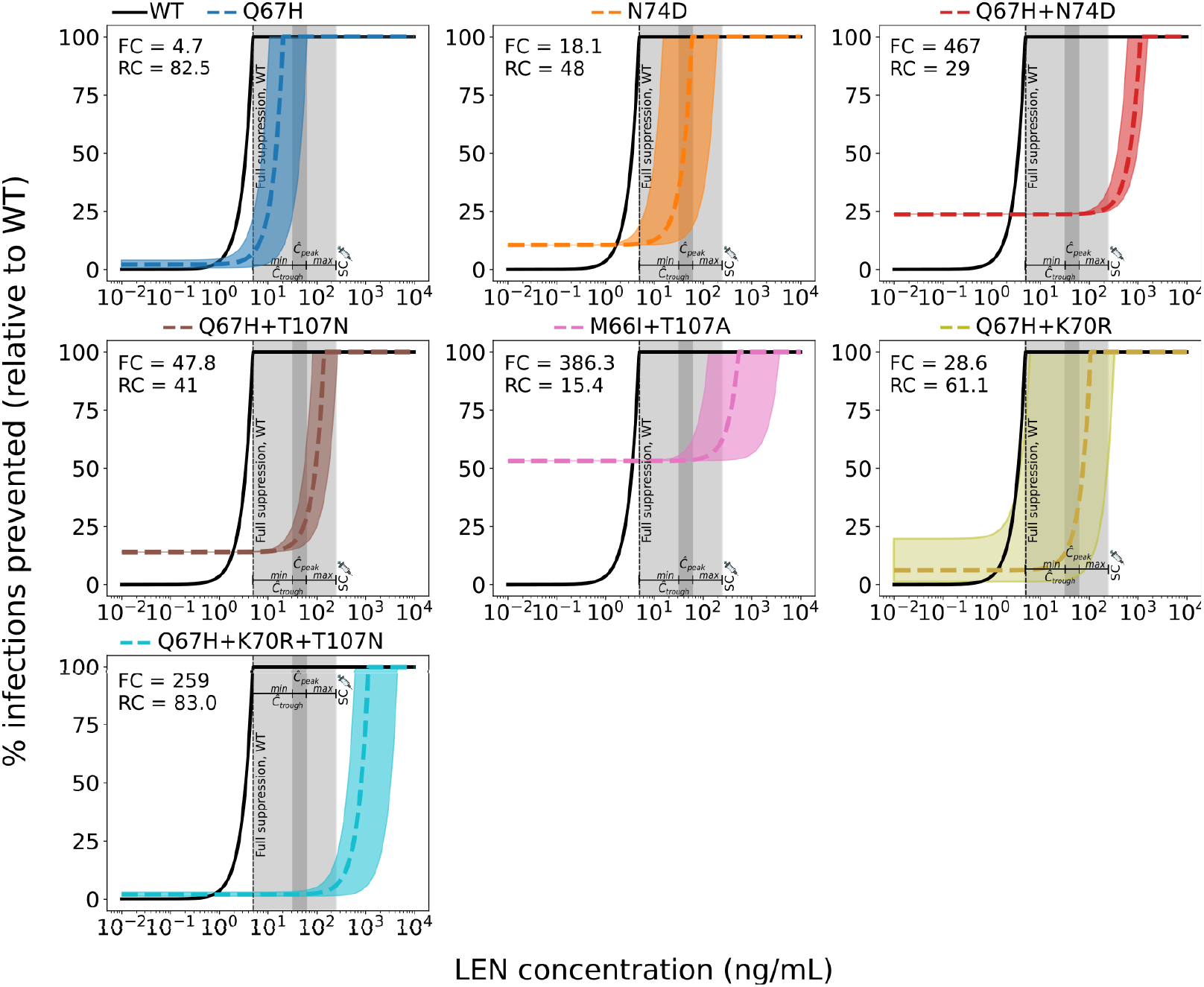
Reduction in HIV infection risk by twice-yearly SC LEN for wild-type and mutant strains. Reduction in HIV wild-type (WT) infection risk (black solid lines) and mutant viruses (colored non-solid lines) for single, double and triple mutants. Variability in mutant-specific fitness and/or fold-change values is shown as colored shaded areas corresponding to each mutation, as labeled at the top of each panel. Infection risk reduction (y-axis) with a particular variant and drug concentrations is computed relative to the infection risk with the WT in the absence of drug. The dark-gray area indicates the steady-state concentration range in an ‘average individual’ (*C*_*trough*_-to-*C*_*peak*_ concentrations) achieved with twice-yearly SC LEN injections (PEG formulation). The light-gray area represent ranges of inter-individual variability in drug concentrations informed by observations in the Purpose 2 trial (*min*-*max*). Complete suppression of WT virus is achieved at a LEN concentration of 5ng/mL (dashed vertical line).

Next, we investigated how LEN-based PrEP with twice-yearly SC administration may facilitate the transmission of drug-resistant viruses (corresponding analysis for once-yearly IM administration is provided in Supplementary Fig. S3). We computed the reduction in HIV infection probability depending on the actual LEN concentration after virus exposure to a mutant strain *i*, relative to wild-type in the absence of drug, eq. (10). This means that in the absence of drug, there may be a reduced HIV risk if a particular mutant strain has an intrinsic fitness disadvantage. At higher LEN concentrations, wild-type infection may become less likely than infections with resistant strains, which increases the *relative proportion* of new infections with transmitted drug resistance.

Fig. 5 illustrates how both the fitness cost and resistance level of each strain affect the overall HIV risk reduction of LEN. Transmission of drug resistance is facilitated whenever the concentration-response curve of the the mutant lies below that of the WT (i.e. mutant transmission becomes more efficient). All investigated mutations (see Supplementary Table S4) show reduced susceptibility to LEN and may compromise the efficacy of PrEP. However, infection with the single mutant Q67H would be completely prevented at population-average steady-state drug levels achieved by SC injections (median prophylactic efficacy of 100%; dark gray area), while the median efficacy may drop to be as low as 9% in some individuals (lower variability limit; light gray area). Infection with the N74D mutant can still occur, but its median infection probability varies between 35–100% in an ‘average individual’, compared with WT in the absence of LEN. Considering inter-individual PK variability (light gray areas in Fig. 5) median reduction of infection risk may be as low as 11% in some individuals, which is solely attributed to intrinsic fitness disadvantage of the N74D mutation, rather than drug efficacy against it. Infection with the double mutants Q67H+T107N and Q67H+K70R cannot be efficiently prevented, showing only a 18-29% and 13-36% median reduction in HIV risk, respectively, across the average steady-state SC concentration range. In contrast, median infection risks associated with the Q67H+N74D (24-27%) and M66I+T107 A (53-63%) double mutants and Q67H+K70R+T107N triple mutant (2-6%) remain almost unaffected, both at average steady-state concentrations and across the full variability range. In summary, these analyses highlight an elevated risk for transmitted drug resistance. For the N74D single mutant, the infection risk varies substantially across concentration ranges typically observed during steady-state subcutaneous administration. An elevated risk was observed for all analyzed double and triple mutants, with Q67H+K70R, Q67H+N74D, Q67H+T107N having high risk and Q67H+K70R+T107N the highest risk for infection with transmitted drug resistance, across the entire concentration range associated with twice-yearly SC-based PrEP.

### De novo emergence of drug resistant variants during LEN PrEP

While transmitted drug resistance increases the *fraction* of incident infections carrying drug resistance mutations, it may not necessarily increase the *absolute number* of individuals infected with resistant virus. I.e., if wild-type infection would be fully prevented whereas infection with resistant virus would not be prevented at all, there would be the same *absolute number* of incident infections with resistant virus under LEN-PrEP. In contrast to transmitted resistance, de novo drug resistance emergence directly elevates the absolute number of individuals carrying drug resistant virus. Herein, we defined de novo drug resistance emerge as an event, where infection with a wild-type virus occurs and where drug resistance is subsequently selected in the newly infected individual. For simulations, we used an approach that considers mutational dynamics and time-varying inhibition of viral replication to compute the probability of infection establishment, compare eqs. (11)-(13).

In particular, we looked at cases where individuals would stop taking LEN injections. In these scenarios, the long pharmacokinetic tail of LEN (Fig. 6A, red line) may give rise to concentrations that allow for infection with wild-type virus and subsequently select drug resistance in the newly infected individual (Fig. 6A, dark blue areas). In our analyses in Fig. 6, we focus on twice-yearly SC injections, while Supplementary Fig. S4 focuses on once-yearly IM injections.

**Fig 6.**
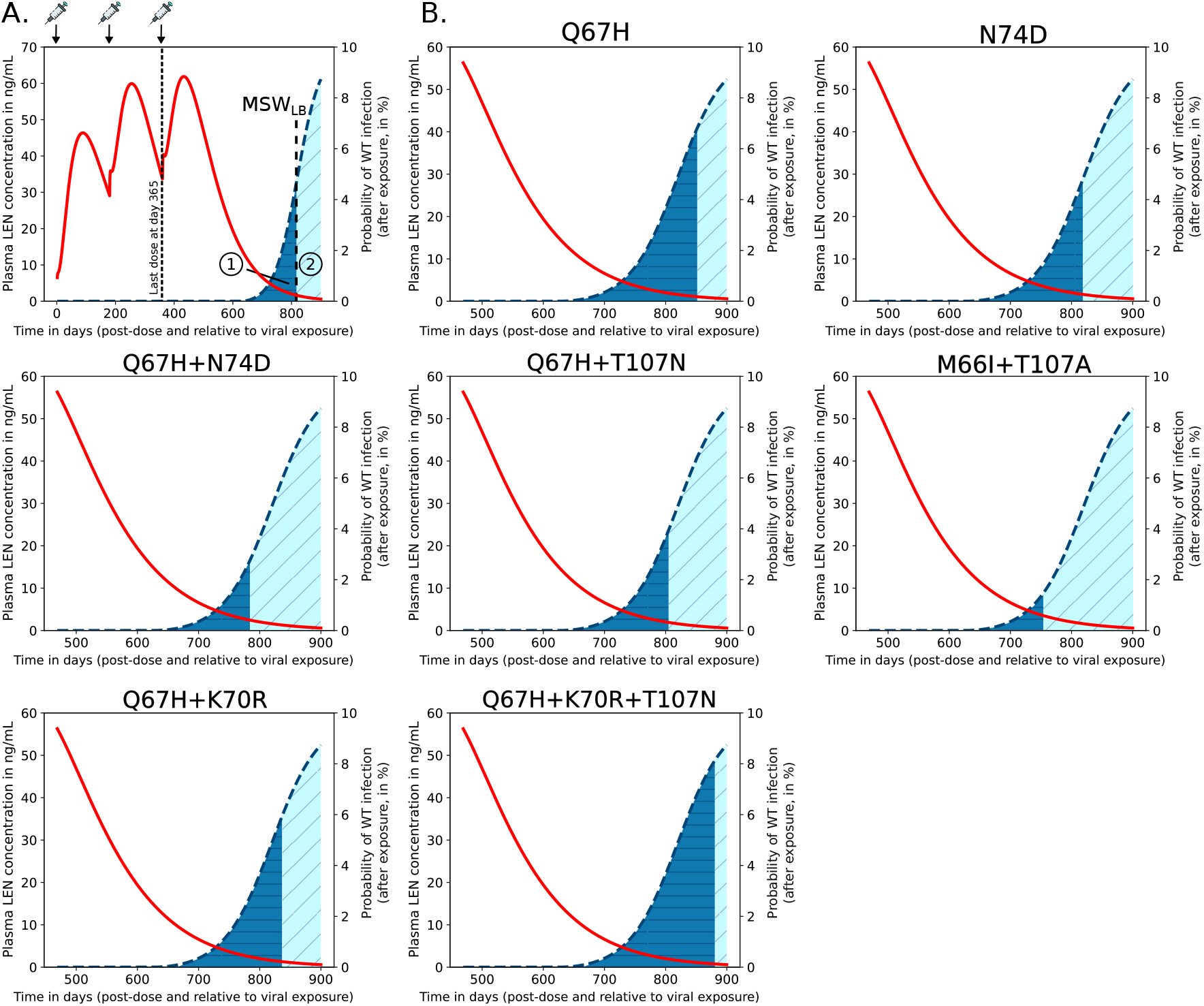
Quantification of de novo drug resistance emergence risk in scenarios where twice-yearly LEN SC doses are missed or when LEN-PrEP is stopped. Predicted average LEN plasma concentrations (red curve; left y-axis) after the first SC LEN injection (PEG formulation) and the probabilities of infection if exposure with WT virus occurred at the indicated time after the last LEN injection (blue dashed curve; right y-axis). A. Example of twice-yearly LEN SC dosing scenario, indicated at the top of the figure. If exposure with WT virus occurs after stopping LEN, two outcomes are possible: (i) infection with WT virus and de novo emergence of a resistant mutant (dark blue area), or (ii) infection with WT virus and selection of WT. The vertical line indicates the lower concentration threshold of the mutant selection window (MSW_*LB*_), i.e. at drug concentrations below this line WT will be selected (see Fig. 4). B. Simulation results for LEN-associated single, double and triple mutants.

For average drug pharmacokinetics observed during twice-yearly SC injections, we predicted that infection with WT virus can occur approximately 285 days (i.e. about 41 weeks) after the last injection. If infection with WT virus occurs at that time, all of the considered drug resistant single-, double- and triple mutants will subsequently be selected. While de novo drug resistance emergence is certain for the indicated times in Fig. 6B (dark blue areas), there is still some residual HIV risk reduction against WT infection. Remarkably, the risk for de novo emergence of drug resistance is considerable during a time window of 206, 170, 138, 160, 106, 191 and 235days for mutations Q67H, N74D, Q67H+N74D, Q67H+T107N, M66I+T107A, Q67H+K70R and Q67H+K70R+T107N. If infection with WT virus occurs after these time windows, resistance is unlikely to emerge (light blue areas). For IM once-yearly dosing, results are shown in Supplementary Fig. S4, indicating that de novo drug resistance emergence may actually happen approximately two years after the last LEN injection.

Next, we wanted to compare the relative likelihood of mutant de novo emerge during the LEN pharmacokinetic tail. We found that Q67H+K70R+T107N was the most likely de novo selected mutation, followed by Q67H, Q67H+K70R, N74D, Q67H+T107N, Q67H+N74D and M66I+T107A. The triple mutant Q67H+K70R+T107N combines high intrinsic fitness with strong resistance, resulting in a pronounced selective advantage under drug pressure in our analyses. However, the de novo emergence of this triple mutant is less likely than that of single mutations, as it would require either the simultaneous occurrence of multiple substitutions or their sequential accumulation. Therefore we suggest that Q67H is more likely to emerge first and may subsequently select +N74D or +T107N. This order was identical for IM injections (Supplementary Fig. S4).

Together these results hint at an important problem: De novo drug resistance emergence with LA-LEN PrEP can happen several months (up to one year for IM dosing) after the last injection, at a time where a former PrEP user may simply not be aware of these risks anymore. Noteworthy, while the moderately resistant single mutant Q67H is more likely to emerge de novo in individuals stopping PrEP, highly resistant strains are more likely to be transmitted to individuals on PrEP (compare Fig. 5 and Fig. 6).

## Discussion

Lenacapavir (LEN), when injected subcutaneously every six months, has shown great promise as long-acting PrEP in the Purpose 1 (cis-gender women) and Purpose 2 (MSM and Transgender women) clinical phase trials [7, 11] and is endorsed by the WHO for HIV pre-exposure prophylaxis. At the time of writing, once-yearly intramuscular injections are evaluated in clinical trials. Long-acting LEN formulations overcome the need for regular pill intake, which appears to be a major barrier to the success of oral PrEP in cis-gender women [12].

For example, the fraction of MSM not taking oral TDF/FTC-based PrEP was only 4–21% in major clinical studies like IPERGAY, HPTN 083, Purpose 2 and DISCOVER [8, 11, 31, 32], whereas the fraction of cis-gender women not taking oral PrEP was 44, 64, 71 and ≈ 90% in HPTN 084, Fem-PrEP, VOICE and Purpose 1 [7, 9, 33, 34]. Factors contributing to adherence differences between MSM and heterosexual women are not completely understood, but disbelieve in PrEP efficacy among cis-gender women [35], as well as stigmatization [36] may constitute barriers to oral PrEP uptake and adherence in women. LEN-based LA-PrEP may help overcome this adherence barrier in cis-gender women, who are most affected by HIV acquisitions globally [6]. However, long-acting agents remain unavailable or cost-prohibitive across much of the globe at the time of writing [37], particularly as major funding programs are being stopped.

An important and insufficiently researched aspect of LA-PrEP are potential risks associated with the emergence of resistant variants, which could limit their clinical scope and relevance for prevention. Notably, LEN has a low mutational barrier to drug resistance, with a single mutation conferring high-level, near-complete insusceptibility [17]. Unlike antiviral treatment which is taken life-long, PrEP denotes a voluntary choice of self-protection, whose (dis-)continuation may be influenced by the availability of insurance coverage, financial means, availability of the products, or perceived risk of HIV infection. Conditioned that PrEP is available, perceived risk denotes a strong predictor of PrEP uptake and continuation [38], but it poorly correlates with actual risk [39].

These observations hint that many individuals stopping PrEP for any of the above mentioned reasons may still be at risk of acquiring HIV. Since LEN-based LA-PrEP may persist years after the last injection, drug concentrations may reside within the mutant selection window for a prolonged period. This constitutes a considerable risk for de novo drug resistance emergence if a person becomes infected after having stopped LEN-based LA-PrEP several months or years prior to virus exposure and infection.

In this work, we developed an integrated PK-PD, viral dynamics and evolution model to improve our understanding of LEN-based PrEP efficacy, and to evaluate the risks of drug resistance development in individuals with a history of LEN-PrEP use. Foremost, we derived a single PK model that allowed to predict oral, SC and IM dosing, for all publicly available, clinically tested single dose LEN formulations. This model was coupled to an established viral dynamics model to predict LEN direct effects from Phase Ib clinical data. Using this integrated model, we were able to predict prophylactic efficacy for all investigated regimens. Further extension of the model with *in vitro* data of clinical isolates allowed to predict the mutant selection window, risks for transmitted drug resistance, as well as the risks for de novo drug resistance emergence in individuals stopping LEN injections.

Considering both population-average steady-state drug concentrations (32-62ng/mL) and the inter-individual variability range (*min*-*max*: 4.8-248ng/mL), our model predicted that twice-yearly SC LEN PrEP remains fully protective against WT infection in the majority of individuals, with a small fraction of individuals not fully protected (Supplementary Fig. S1). Furthermore, we observed limited protection against certain drug-resistant variants. Notably, two individuals in the LEN arm of the Purpose 2 study, both with sexually transmitted infections (STI, Syphillis, Clamydia), acquired HIV infection [11]. According to the analysis conducted in the Purpose 2 study, both infections may have occurred shortly after initiating LEN, possibly during the lead-in (i.e. pre-steady-state) phase. Interestingly, both individuals had viruses carrying drug resistance with the N74D mutation, which confers strong resistance *in vitro* [20] and is selected during LEN treatment [40]. While N74D alone implies a fitness deficit based on clinical isolate data, we predicted it to be selected at a wide range of drug concentrations (1.7-59 ng/mL in Fig. 4). However, fitness deficits may in vivo be overcome by compensatory mutations [20], which may lead to their persistence, once selected. Notably, the combination Q67H+N74D confers strong resistance, but has some fitness deficit (infection probability reduced by 75% in comparison to WT; Fig. 5), according to *in vitro* data. Clinically, N74D has been observed more frequently with M66I, while Q67H has been frequently observed together with K70R [20], which have almost WT-like fitness and can also be transmitted at clinically relevant concentrations (Fig.5).

In one infected individual in Purpose 2, LEN concentrations were consistently *>* 20ng/mL at week 4–13; -concentrations at which WT infection is unlikely (compare Fig. 5), arguing that infection may either have occurred at pre-steady-state in this individual or with transmitted drug resistance. The other individual infected in Purpose 2 had consistently lower than average concentrations of LEN (5–20ng/mL at weeks 4–26). According to our predictions, either (i) infection with WT occurred during the pre-steady-state phase, or (ii) infection with WT virus occurred at steady-state in this individual, followed by subsequent resistance selection, (iii) or resistance was transmitted. The dynamics of drug resistance emergence, in case of HIV infection occurs shortly before LEN application, or in the pre-steady-state phase may be similar to those observed during monotherapy, with distinct possible evolutionary trajectories involving any of the analyzed mutations alone or in combination [20]. Another possibility is that inter-individual variability with regards to within-host antiviral mechanisms (innate and adaptive immunity) may also contribute to variability in preventive plasma concentrations (e.g. *EC*_95_; fully preventive concentrations), warranting further investigations.

Notably, resistance-associated mutations in the HIV capsid may be low in frequency in consensus sequences [41], but they are not absent in minority variants [42] and the HIV-1 capsid appears to be intrinsically variable [43]. Therefore, the occurrence of transmitted drug resistance may be low, but cannot be entirely ruled out (hypothesis iii). Infection with WT virus at steady-state may be unlikely (hypothesis ii), but also not impossible for the second individual.

A major limitation of our analysis is that due to the limited availability of individual data (see Supplementary Table S1), we were not able to to fit and parameterize population pharmacokinetic models. To approximate inter-individual pharmacokinetic variability, we incorporated summary-level PK variability from the Purpose 2 trial (see Supplementary Fig. S1), which highlighted that plasma concentrations may vary by approximately one order of magnitude between individuals. While this approach provides a reasonable estimate of concentration ranges, it cannot substitute for comprehensive Population-PK modeling. Nevertheless, our approach highlights that infections, albeit with small probability, may occur at steady-state twice-yearly LEN injections.

In our work, we predicted that the risk for de novo drug resistance emergence after wild-type infection starts to rise when concentrations fall below 5ng/mL, which occurs approximately 11.8 months after the last injection of 927mg SC in an “average” individual, but may also occasionally be encountered during steady-state twice-yearly dosing, according to [11]. Based on utilized viral fitness parameters, we found that Q67H is the most likely mutation to be selected in this scenario (due to its relatively high fitness). Interestingly, Q67H emerged in 2 participants receiving low SC doses (20mg and 50mg LEN) in a Phase Ib treatment study [44], which would be consistent with our predictions (Fig. 6) indicating that the Q67H mutation is preferentially selected at low LEN concentrations (*>* 0.8 ng/mL, Fig. 4). Noteworthy, while we predicted that the single mutant Q67H is more likely to emerge de novo in individuals stopping PrEP, it is not likely to be transmitted to individuals on PrEP (compare Fig. 5 and Fig. 6). I.e. transmitted drug resistance may favor strongly resistant variants, whereas de novo selection favors variants that confer little fitness cost.

Our simulations are based on mutant phenotype data (i.e. selective disadvantage and fold-change in *IC*_50_) obtained from *in vitro* experiments of clinical isolates. However, mutation-specific parameters derived *in vitro* may not fully reflect in vivo viral fitness or resistance dynamics. In vivo, mutant phenotypes may be influenced by the presence or pre-existence of compensatory mutations through epistatic interactions [45], partly explaining differences between *in vitro* and in vivo derived phenotypes [20], and potentially explaining distinct evolutionary trajectories of LEN failure through drug resistance development. For mathematical modeling, it is impossible to anticipate the different genetic backgrounds of susceptible viruses, which may subsequently favour one-over another evolutionary trajectory to drug resistance. Consequently, the utilized phenotypic parameters may not fully account for the fitness of virus variants carrying the indicated mutations and, in turn, may not fully capture the risk of de novo resistance emergence.

Nonetheless, our results hint at an important problem: De novo drug resistance emergence with LA-LEN PrEP can happen several months (or years) after the last injection, at a time when a former PrEP user may not be aware of these risks anymore. These findings argue for public health considerations of the risks and benefits of LEN-based LA-PrEP and the need to develop LEN-PrEP discontinuation strategies.

Notably, the risk of de novo drug resistance emergence after PrEP discontinuation may similarly exist for long-acting cabotegravir, which also has a low barrier to drug resistance and which may persist month after discontinuation. In contrast, islatravir constitutes a very large genetic barrier to resistance and drug concentrations quickly decay after removal of ISL implants [10], which would minimize those risks.

Unlike long-acting PrEP, the risk of de novo drug resistance emergence during oral TDF/FTC-based PrEP may arise by different adherence mechanisms. The risk for resistance emergence after terminally stopping oral PrEP may be extremely low because of a high genetic barrier to drug resistance, as well as comparably short drug half-lives (i.e. ≈ 1.6 days for FTC-TP and ≈ 6.5 days for TFV-DP) [46]. These shorter drug half-lives may simply not provide enough “window of opportunity” for WT infection to subsequently select de novo drug resistance. Low constant levels of adherence, or frequent dis- and re-continuation may however create environments in which infection with WT can occur with subsequent drug resistance selection when oral TDF/FTC-based PrEP is mistakenly re-initiated after undetected infection (i.e. as monotherapy) [47].

### Study highlights

#### What is the current knowledge on the topic?

Twice-yearly long-acting lenacapavir(LEN)-PrEP has completed Phase III clinical trials demonstrating *>*90% HIV risk reduction and was approved by the FDA (July 2025), with subsequent endorsement by the WHO for HIV PrEP.

#### What question did this study address?

Key aspects of LEN-PrEP remain unclear: (i) Preventive concentration benchmarks and prophylactic efficacy is challenging to quantify from clinical studies and (ii) LEN persists in individuals stopping PrEP for over a year, posing a risk of de novo drug resistance development.

#### What this study adds to our knowledge?

We developed an integrated PK/PD model of LEN, incorporating PK variability, to evaluate efficacy against wild-type and transmitted drug-resistant HIV and to assess the risk of de novo resistance after discontinuation of LEN-PrEP.

#### How might this change drug discovery, development, and/or therapeutics?

Twice-yearly LEN represents a breakthrough in HIV PrEP. Our study informs the development of formulations that achieve protective concentrations, supports evaluation of once-yearly IM dosing, and underscores the importance of discontinuation strategies to mitigate de novo resistance in the context of LEN-PrEP.

## Data Availability

Zenodo, GitHub

https://doi.org/10.5281/zenodo.16612337

https://github.com/KleistLab/LenPrEP

## Data and code availability

All analysis were performed using custom codes written in Python 3.12 and are available at https://github.com/KleistLab/LenPrEP under GPL 3.0 open access license. A frozen version of the code to reproduce all findings is available at Zenodo (https://doi.org/10.5281/zenodo.16612337).

## Acknowledgments

H-Y.K. and M.v.K. acknowledge funding from the German Federal Ministry of Research, Technology and Space (BMFTR; grant number 01KI2016). A.L. acknowledges an internal grant for PhD projects provided through the Robert Koch-Institute. L.Z. acknowledges funding from the German Ministry for Health (BMG; grant number: ZMII2-2524FSB62A) and M.v.K. acknowledges funding by the Deutsche Forschungsgemeinschaft (DFG, German Research Foundation) under Germany’s Excellence Strategy – The Berlin Mathematics Research Center MATH+ (EXC-2046/1, project ID: 390685689). The funders had no role in designing the research or the decision to publish.

## Conflict of interest statement

All other authors declared no competing interests for this work.

## Author Contributions

H-Y.K. and M.v.K. wrote the manuscript with help from L.Z and A.L. H-Y.K. and M.v.K. designed the research. H-Y.K., A.L and L.Z. performed the research and H-Y.K. and M.v.K analyzed the data.

## Notes

### Competing Interest Statement

The authors have declared no competing interest.

### Funding Statement

HY.K. and M.v.K. acknowledge funding from the German Federal Ministry of Research, Technology and Space (BMFTR). A.L. acknowledges an internal grant for PhD projects provided through the Robert Koch-Institute. L.Z. acknowledges funding from the German Ministry for Health (BMG) and M.v.K. acknowledges funding by the Deutsche Forschungsgemeinschaft (DFG, German Research Foundation) under Germany's Excellence Strategy, The Berlin Mathematics Research Center MATH+ (EXC-2046/1). The funders had no role in designing the research or the decision to publish.

### Summary of Updates

We revised the PK model and, based on a recent study, incorporated mutation-specific parameters derived from in vitro phenotypic assays of clinical isolates, including reported variability estimates where available. We updated all corresponding simulations accordingly.

