## Supporting Text for "Pharmacokinetics, pharmacodynamics, efficacy and drug resistance selection of injectable long-acting lenacapavir pre-exposure prophylaxis (PrEP) against HIV"

##### Overview clinical PK data

| Study ID [Ref.] | Study design/Focus | Formulation | Dataset |
|---|---|---|---|
| GS-US-200-4070 [1,2] (Study 1) | Phase I in healthy subjects ($n = 8$/cohort). Safety, tolerability and SC PK. | SC aqueous suspension | 30, 100 mg, 300 and 450 mg |
| GS-US-200-4071 [3] (Study 2) | Phase I in healthy subjects ($n = 8$/cohort). Effect of food on oral PK. | Oral tablets | 300, 900 and 1,800 mg |
| GS-US-200-4072 [1] (Study 3) | Phase Ib in HIV-1 infected subjects ($n = 6$/cohort). Antiviral activity. | SC aqueous suspension | 20 mg, 50 mg, 150 and 450 mg |
| GS-US-200-4358 [3,4] (Study 4) | Phase I in healthy subjects ($n = 8$/cohort). Safety, tolerability and SC PK. | SC PEG/water solution | 309 mg, 927 mg |
| Study 5 [5] | Phase I in healthy subjects ($n = 20$/cohort). Safety, tolerability and IM PK. | IM ethanol/ water solution | 5000 mg with 5% w/w ethanol (F1), 10% w/w ethanol (F2) |

**Table S1. Summary table of publicly available PK/PD data used for LEN.** In total, 15 datasets were analyzed: 3 for oral administration, 10 for subcutaneous (SC), and 2 for intramuscular (IM) injections. All parenteral formulations showed distinct kinetics and were modeled separately.

#### Clinical and model-based PK properties

| ID | Dose (in mg) | $C_{max}$ (ng/mL) | | $T_{max}$ (days) | | $t_{1/2}$ (days) | |
|---|---|---|---|---|---|---|---|
| | | study | model | study | model | study | model |
| SC aq. (Study 1) | 30 | 3.2 ± 1.3 | 3.1 | 35.0 ± 7.3 | 20.9 | 35.5 ± 4.4 | 45.9 |
| | 100 | 14.7 ± 8.6 | 10.2 | 21.0 ± 12.5 | 21 | 30.3 ± 11.8 | 45.9 |
| | 300 | 47.9 ± 13.3 | 30.7 | 31.5 ± 8.8 | 20.9 | 43.1 ± 10 | 45.9 |
| | 450 | 58.4 ± 13.4 | 46.1 | 14.0 ± 11.7 | 20.8 | 39.9 ± 8 | 45.9 |
| Oral (Study 2) | 300 | 33.7 ± 32.5 | 21.3 | 0.2 ± 0.04 | 0.2 | 11 ± 2.6 | 11.1 |
| | 900 | 43.9 ± 32.2 | 24.1 | 0.2 ± 0.4 | 0.2 | 13.4 ± 2 | 11.1 |
| | 1,800 | 53.8 ± 25.8 | 34.3 | 0.3 ± 0.06 | 0.2 | NA | 11.1 |
| SC aq. (Study 3) | 20 | 2.5† | 2.1 | 9.0† | 9 | 0.7† | |
| | 50 | 4.3† | 5.2 | 7.0† | 9 | 0.7† | |
| | 150 | 15.0† | 15.7 | 6.0† | 9 | 0.7† | |
| | 450 | 43.7† | 47.1 | 8.0† | 9 | 0.7† | |
| SC PEG (Study 4) | 309 | 17.7 ± 8.9 | 16.1 | 98 ± 42.1 | 84 | 106 ± 43.2 | 65 |
| | 927 | 67.0 ± 36.7 | 48.2 | 77 ± 7.1 | 84 | NA | 65 |
| IM (Study 5) | 5000 (F1) | 247.0 ± 81 | 176.2 | 84.1 ± 28.0 | 112.5 | NA | 106.8 |
| | 5000 (F2) | 336.0 ± 120.4 | 219.8 | 69.9 ± 25.1 | 83.9 | NA | 106.8 |

**Table S2. Overview of clinical PK properties and model-based calculations for oral, subcutaneous (SC) and intramuscular (IM) formulations.** The reported PK values comprise $C_{max}$ (maximal concentration), $T_{max}$ (time to reach $C_{max}$) and $t_{1/2}$ (half-life). Values are taken from published clinical trial results or study protocols (GS-US-200-4334, GS-US-200-4072). Model-based calculations of the PK values are shown alongside the reported data. The values represent the mean (± std) of the considered doses, except for $T_{max}$ and $t_{1/2}$, which are reported as the median (± Q3-Q1/2) with Q1 = first quartile and Q3 = thrid quartile. If clinical parameters were not available, PK values extracted from the data are reported and marked with a superscript $^{\dagger}$, otherwise, they are marked as NA (not available). Values that fall outside the reported range are shown in gray.

#### Estimation of inter-individual variability in 927 mg SC LEN PrEP

To incorporate pharmacokinetic variability into our analysis, we digitized the published median, minimum, and maximum concentration profiles of the Purpose 2 study and expressed the lower and upper bounds as ratios relative to the median at each time point [6]. These concentration–time profiles represent a 10% subsample of participants who received 927 mg SC LEN, including two individuals in the LEN PK group who acquired HIV infection. Plasma concentrations were reported from week 4 to 52 (6 time points), covering two SC injections.

Since there were only minor differences between median and mean values (compare Supplementary Fig. S1), the resulting scaling factors were applied to the 927 mg SC regimen (Study 4), which reported average concentrations following single-dose administration. Missing values (i.e., absorption and tail phases) in both concentration–time profiles were imputed by interpolation or extrapolation. To summarize the variability, we calculated the geometric mean of the factors, yielding 0.15 for the lower and 4.0 for the upper concentration limits, which we subsequently applied to the observed steady-state concentration range. Variability factors were also applied to other model simulations.

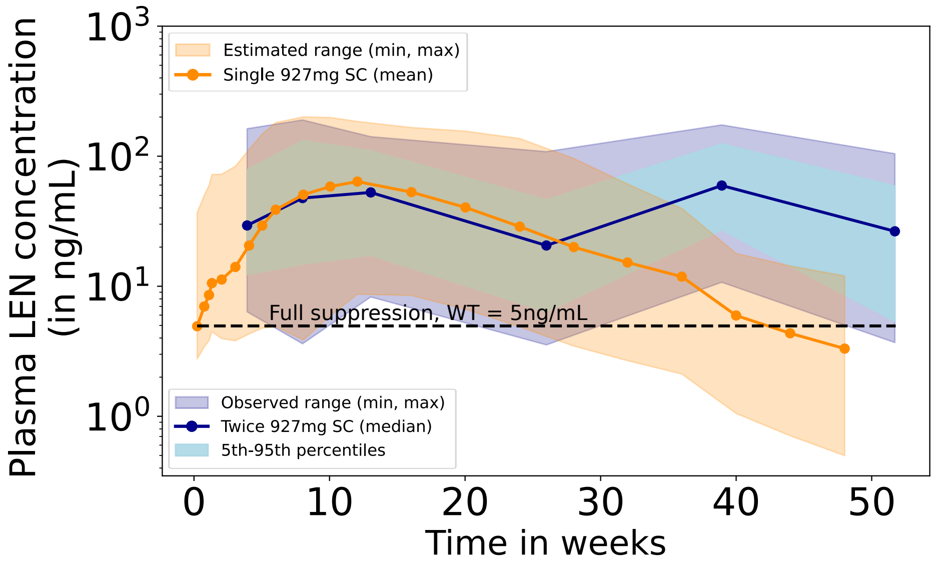


**Figure S1. Inter-individual variability in LEN plasma concentrations after 927 mg SC administration.** The blue line shows median concentrations in the pharmacokinetics cohort, with the light-blue shaded area indicating the 5th–95th percentiles and the dark-blue shaded area representing the minimum–maximum range observed in the Purpose 2 trial (twice-yearly SC dosing; dosing events at weeks 0 and 26). The orange line represents PK data after a single 927 mg SC dose (Study 4), including mean, minimum, and maximum concentrations, with imputed variability (derived by interpolation and extrapolation). A dashed horizontal line marks the concentration threshold for complete suppression of WT virus (5ng/mL LEN).

#### PK-viral dynamics model for estimating LEN potency

The decline in viral load (i.e. the inferred antiviral effect $IC_{50}$), following LEN monotherapy was modeled by linking the pharmacokinetics model to an established HIV-1 viral dynamics model comprising free infectious- and uninfectious virus $V$ and $V_N$, as well as uninfected, early and late infected T-cells and macrophages ($T_u$, $T_1$, $T_2$ and $M_u$, $M_1$, $M_2$). The corresponding system of ODEs and parameter values are given below [7, 8].

$$\frac{d}{dt}T_u = \lambda_T - \delta_T \cdot T_u - \beta_T \cdot V \cdot T_u + \delta_{\text{PIC,T}} \cdot T_1 \tag{S1}$$

$$\frac{d}{dt}M_u = \lambda_M - \delta_M \cdot M_u - \beta_M \cdot V \cdot M_u + \delta_{\text{PIC,M}} \cdot M_1 \tag{S2}$$

$$\frac{d}{dt}T_1 = \beta_T \cdot V \cdot T_u - (\delta_{T1} + k_T + \delta_{\text{PIC,T}}) \cdot T_1 \tag{S3}$$

$$\frac{d}{dt}M_1 = \beta_M \cdot V \cdot M_u - (\delta_{M1} + k_M + \delta_{\text{PIC,M}}) \cdot M_1 \tag{S4}$$

$$\frac{d}{dt}T_2 = k_T \cdot T_1 - \delta_{T2} \cdot T_2 \tag{S5}$$

$$\frac{d}{dt}M_2 = k_M \cdot M_1 - \delta_{M2} \cdot M_2 \tag{S6}$$

$$\frac{d}{dt}V = N_T \cdot T_2 + N_M \cdot M_2 - V \cdot (CL + (\beta_T + CL_T) \cdot T_u + (\beta_M + CL_M) \cdot M_u) \tag{S7}$$

$$\frac{d}{dt}V_N = \left(\hat{N}_T - N_T\right) \cdot T_2 + \left(\hat{N}_M - N_M\right) \cdot M_2 - CL \cdot V_N \tag{S8}$$

| **T cells** | | **Macrophages** | |
|---|---|---|---|
| Parameter | Value | Parameter | Value |
| $\lambda_T$ | $2 \times 10^9$ | $\lambda_M$ | $6.9 \times 10^7$ |
| $\delta_T$ | 0.02 | $\delta_M$ | 0.0069 |
| $\delta_{T1}$ | 0.7 | $\delta_{M1}$ | 0.0069 |
| $\delta_{T2}$ | 1 | $\delta_{M2}$ | 0.09 |
| $\delta_{\text{PICT}}$ | 0.35 | $\delta_{\text{PICM}}$ | 0.0035 |
| $k_T$ | 0.35 | $k_M$ | 0.07 |
| $\beta_T$ | $1.8 \times 10^{-12}$ | $\beta_M$ | $2 \times 10^{-14}$ |
| $\hat{N}_T$ | 1000 | $\hat{N}_M$ | 100 |
| $N_T$ | 670 | $N_M$ | 67 |

**Table S3.** Parameters used for the viral dynamics model were adapted from [7, 8] (original sources indicated therein). All parameters refer to the absence of any drug. Production rates $\lambda_T$ and $\lambda_M$ are given in cells/day, and infection rate constants $\beta_T$ and $\beta_M$ (lumped rates of successful target cell infection) in 1/(day·virus). All remaining parameters are given in 1/day. The parameters $\delta$ denote cellular death rate constants, whereas $\delta_{\text{PIC},T}$ and $\delta_{\text{PIC},M}$ describe intracellular degradation of the pre-integration complex leading to the reversion to uninfected cells. The parameters $k_T$ and $k_M$ denote viral genome integration rate constants, and $\hat{N}_{T/M}$ and $N_{T/M}$ the total and infectious virus production rates from late infected cells, respectively. The viral kinetic parameters, with $\beta_T = 1.8 \times 10^{-12}$ and $\beta_M = 2 \times 10^{-14}$, were adjusted to reproduce the patients' baseline viral load of approximately 4.5 $\log_{10}$ HIV-1 RNA copies/mL in plasma. The clearance rate was fixed to $CL = 23$ (1/day) in infected individuals (simulation of treatment effects) [7] and $CL = 2.3$ (1/day) in virus-naive individuals (PrEP simulations) [9]. Parameter $CL_{T/M} = (1/0.5 - 1) \cdot \beta_{T/M}$ in 1/(day ·virus) [9]. The estimated pharmacodynamic parameter was $IC_{50} = 1.6$ ng/mL (see *Methods*).

#### Phenotypic susceptibility (FC) and viral fitness (RC) of LEN capsid resistance mutations

Phenotypic mutation parameters were derived from *in vitro* characterization of clinical isolates and are reported as median or mean values with variability when available. *In vitro* resistance selection experiments identified seven major LEN-associated capsid mutations (L56I, M66I, Q67H, K70N, N74D, N74S, T107N) and their combinations [10].

| | $EC_{50}$ fold-change (FC) | Replication capacity (relative fitness %) | |
|---|---|---|---|
| Resistance mutation | Median (min–max) | Average (min–max) | Ref. |
| Q67H | 4.7 (2.7–13.8) | 82.5 (71, 94) | [10] |
| N74D | 18.1 (4.7–59.7) | 48 | [10, 11] |
| Q67H+N74D | 467 (263.4–670.5) | 29 | [10, 11] |
| Q67H+T107N | 47.8 (27.9–89.2) | 41 | [10, 11] |
| M66I+T107A | 386.3 (94.3–2700) | 15.4 | [10] |
| Q67H+K70R | 28.6 (2.4–72.5) | 61.15 (33, 89.3) | [10] |
| Q67H+K70R+T107N | 259 (145–1000) | 83.05 (76.4, 89.7) | [10] |

**Table S4. Treatment-emergent HIV-1 capsid resistance mutations and associated phenotypic characteristics.** Lenacapavir (LEN) fold change (FC) values (unitless) represent the change in $EC_{50}$ relative to wild-type (WT) virus and are reported as median (minimum–maximum) values derived from clinical isolates [10]. Replication capacity (relative fitness $s(i)$ in %) of HIV-1 variants carrying LEN-resistance mutations compared to WT virus is reported as the average if multiple measurements were available (ranges), or as single data points when only one measurement was available. Replication capacity values were obtained from [10] or [11], as indicated.

### Viral extinction and infection probability

We utilized a simplified viral dynamics model (no macrophage cells and no non-infectious virus) in comparison to the model used to predict viral load decay (section *PK-viral dynamics model for estimating LEN potency*), as previous work [12, 13] indicated that the simpler model is sufficient to predict PrEP efficacy. Furthermore, in a PrEP scenario, we can assume that target cells are approximately constant levels at $T_u = \lambda_T/\delta_{T_u}$, since infection, which would decrease their levels, has not yet happened. This reduced model may also include mutational dynamics, particularly when we study *de novo* emergence of drug resistance.

$$\frac{d}{dt}T_1(i) = \sum_j V(j) \cdot p_{j\to i} \cdot \beta_T \cdot T_u - T_1(i) \cdot \big(\delta_{T_1} + \delta_{PIC} + k_T\big) \tag{S9}$$

$$\frac{d}{dt}T_2(i) = k_T \cdot T_1(i) - T_2(i) \cdot \delta_{T_2} \tag{S10}$$

$$\frac{d}{dt}V_I(i) = s(i) \cdot \Big(1 - \eta\big(C_t, FC(i)\big)\Big) \cdot N_T \cdot T_2(i) - V(i) \cdot \Big(CL + \big(CL_T + \beta_T\big) \cdot T_u\Big), \tag{S11}$$

where $V$ denotes infectious virus and parameters as defined above. Here, the selective disadvantage of a mutant strain $i$, $s(i) \leq s(WT) = 1$ (defined as 1 - relative fitness), reflects the loss in replication ability of a resistant phenotype relative to the wild-type virus and $FC(i) \geq 1$ denotes a fold change in drug susceptibility of the mutant virus $i$. These values were derived from *in vitro* phenotypic assays of clinical isolates, incorporating variability in fold-change and intrinsic fitness values (details in Supplementary Table S4). The probability that any strain $j$ mutates into another strain $i$ was given by the transition probability $p_{j\to i}$ and accounts for all possible states along the mutagenic pathway. We computed the transition probability based on the Hamming distance $h(i,j)$ between strains $i$ and $j$ in terms of the number of amino acid substitutions required to convert one genotype into the other. Assuming that each position mutates independently, the transition probability is calculated as:

$$p_{j\to i} = \mu^{h(i,j)} \cdot (1-\mu)^{N-h(i,j)} \tag{S12}$$

where $\mu$ represents the mutation probability per base ($\mu \approx 2.16 \cdot 10^{-5}$ [7]), and $N$ denotes the total number of mutated positions.

For notational ease, we compile reaction rates into six reactions classes $R_1$ to $R_6$ based on their shared stoichiometry and derive reaction propensities $a_1$-$a_6$:

$$R_1: \text{Clearance of free virus,} \quad V \to \varnothing \quad a_1 = (CL + CL_T \cdot T_u) \cdot V \tag{S13}$$

$$R_2: \text{Clearance of } T_1\text{-cell,} \quad T_1 \to \varnothing \quad a_2 = (\delta_{PIC} + \delta_{T_1}) \cdot T_1 \tag{S14}$$

$$R_3: \text{Clearance of } T_2\text{-cell,} \quad T_2 \to \varnothing \quad a_3 = \delta_{T_2} \cdot T_2 \tag{S15}$$

$$R_4: \text{Infection of a suscept. cell,} \quad V \to T_1 \quad a_4 = \beta \cdot T_u \cdot V \tag{S16}$$

$$R_5: \text{Integration of viral DNA,} \quad T_1 \to T_2 \quad a_5 = k \cdot T_1 \tag{S17}$$

$$R_6: \text{Production of new virus,} \quad T_2 \to V + T_2 \quad a_6(t,i) = s(i) \cdot (1 - \eta(C_t, i))\, N_T \cdot T_2, \tag{S18}$$

where $(1 - \eta(C_t, FC(i)))$ denotes the (time-dependent) inhibition of virus production against strain $i$ by LEN, as outlined in the main manuscript and $s(i)$ denotes some intrinsic fitness disadvantage of a mutant $i$ with $s(i) \leq s(WT) = 1$.

The model is sketched in Fig. 2, main text.

#### Analytical solution for infection probability (constant drug, no mutational dynamics)

For the model above, we define a multi-type branching process with states $V$, $T_1$ and $T_2$ and reaction rates $a_1$ to $a_6$, in analogy to [9]. We assume (i) constant drug concentrations and (ii) no mutational dynamics. This analytical solution is only used for mutant window analysis and analysis of transmitted drug resistance. Assuming statistical independence $P_E(n \cdot \hat{V}) = \big(P_E(\hat{V})\big)^n$, for a virus variant $i$ (e.g. 'WT', 'Q67H', ...) the extinction probability $P_E(\hat{V})$ after exposure to a single virus particle $\hat{V} = [1, 0, 0]$ is then given by:

$$P_E(Y_0 = \hat{V}) = \sum_{n=0}^{\infty} \mathbb{P}(Y_r = n \cdot \hat{V} | Y_0 = \hat{V}) \cdot \big(P_E(Y_r = \hat{V})\big)^n \tag{S19}$$

$$\Leftrightarrow P_E(\hat{V}) = \sum_{n=0}^{\infty} \mathbb{P}(Y_r = n \cdot \hat{V} | Y_0 = \hat{V}) \cdot \big(P_E(\hat{V})\big)^n \tag{S20}$$

where we used that $P_E(Y_0 = \hat{V}) = P_E(Y_r = \hat{V}) = P_E(\hat{V})$. Above, $\mathbb{P}(Y_r = n \cdot \hat{V} | Y_0 = \hat{V})$ denotes the probability that $n$ viruses emerged from a single replication cycle $r$ conditioned on an initial state of one virus, $Y_0 = \hat{V}$. Let us introduce the following transition probabilities:

$$p_1 = p_{V \to \varnothing | V} = \frac{a_1}{a_1 + a_4} \qquad p_2 = p_{T_1 \to \varnothing | T_1} = \frac{a_2}{a_2 + a_5} \qquad p_3(i) = p_{T_2 \to \varnothing | T_2}(i) = \frac{a_3}{a_3 + a_6(i)} \tag{S21}$$

i.e. the probability of clearance, conditioned the system is in states $V$, $T_1$ and $T_2$ respectively. Note that reaction propensity $a_6(i)$ in this framework can be defined considering a constant drug inhibition, fold resistance and selective disadvantage, i.e. $a_6(i) = s(i) \cdot \big(1 - \eta(C, FC(i))\big) \cdot a_6(WT, \varnothing)$, where $a_6(WT, \varnothing)$ denotes the reaction propensity for infectious virus production in the wild-type in the absence of LEN and $C$ and $FC(i)$ denote a (static) drug concentration and a fold change for mutant $i$.

From here, we can define the remaining transition probabilities, $(1 - p_1) = p_{V \to T_1 | V}$, $(1 - p_2) = p_{T_1 \to T_2 | T_1}$ and $(1 - p_3(i)) = p_{T_2 \to T_2 + V | T_2}$. We then write out the first term in eq. (S20) for $n = 0$ as $\mathbb{P}(Y_r = 0 \cdot \hat{V} | Y_0 = \hat{V}) = p_1 + (1 - p_1) \cdot p_2 + (1 - p_1)(1 - p_2) \cdot p_3(i)$, as well as the terms for $n > 0$ as $\mathbb{P}(Y_r = n \cdot \hat{V} | Y_0 = \hat{V}) = (1 - p_1)(1 - p_2)(1 - p_3(i))^n \cdot p_3(i)$. Substituting and simplifying yields

$$\begin{aligned} P_E(\hat{V}) &= p_1 + (1 - p_1) \cdot p_2 + (1 - p_1)(1 - p_2) \cdot p_3(i) \cdot \sum_{n=0}^{\infty} \Big((1 - p_3(i)) \cdot P_E(\hat{V})\Big)^n \\ &= p_1 + (1 - p_1) \cdot p_2 + (1 - p_1)(1 - p_2) \cdot p_3(i) \frac{1}{1 - (1 - p_3(i)) \cdot P_E(\hat{V})} \end{aligned} \tag{S22}$$

by solving the geometric series and yielding the following quadratic problem

$$\begin{aligned} 0 =& - P_E(\hat{V})^2 (1 - p_3(i)) + P_E(\hat{V}) \Big(1 + p_1(1 - p_3(i)) + (1 - p_1)p_2(1 - p_3(i))\Big) \\ & - \Big(p_1 + (1 - p_1)p_2 + (1 - p_1)(1 - p_2)p_3(i)\Big) \\ =& P_E(\hat{V})^2 (1 - p_3(i)) - P_E(\hat{V}) \Big(1 + (1 - p_3(i))\big(p_1 + (1 - p_1)p_2\big)\Big) \\ & + \Big(1 + (1 - p_3(i))\big(p_1 + (1 - p_1)p_2\big) - (1 - p_3(i))\Big) \\ =& P_E(\hat{V})^2 - P_E(\hat{V}) \Big(\frac{1}{(1 - p_3(i))} + \big(p_1 + (1 - p_1)p_2\big)\Big) \\ & + \Big(\frac{1}{(1 - p_3(i))} + \big(p_1 + (1 - p_1)p_2\big) - 1\Big) \end{aligned} \tag{S23}$$

where we re-wrote the last term. The quadratic problem yields solution

$$P_E^{(1,2)}(\hat{V}) = \frac{1}{2}\Big(\frac{1}{(1 - p_3(i))} + \big(p_1 + (1 - p_1)p_2\big)\Big) \pm \frac{1}{2}\Big(\frac{1}{(1 - p_3(i))} + \big(p_1 + (1 - p_1)p_2\big) - 2\Big). \tag{S24}$$

The first solution yields $P_E^{(1)}(\hat{V}) = 1$ (certain extinction), whereas the second solution yields

$$P_E^{(2)}(\hat{V}) = \left(\frac{1}{(1-p_3(i))} + \big(p_1 + (1-p_1)p_2\big)\right) - 1 = p_1 + (1-p_1)\cdot p_2 + \frac{p_3(i)}{1-p_3(i)}. \tag{S25}$$

Hence, the extinction probability is

$$P_E(\hat{V}) = \min\left(1,\ p_1 + (1-p_1)\cdot p_2 + \frac{p_3(i)}{1-p_3(i)}\right) \tag{S26}$$

The infection probability is $P_{inf}(\hat{V}) = 1 - P_E(\hat{V})$ and using $1 - \min(1,x) = \max(0, 1-x)$ we get

$$\begin{aligned}
P_{inf}(\hat{V}) &= \max\left(0, (1-p_1)(1-p_2) - \frac{p_3(i)}{1-p_3(i)}\right) \\
&= \max\left(0, \frac{a_4}{a_1+a_4}\cdot\frac{a_5}{a_2+a_5} - \frac{a_3}{a_6(i)}\right) \\
&= \max\left(0, \Lambda - \frac{a_3}{a_6(i)}\right) = \max\left(0, \Lambda\left(1 - \frac{1}{\Lambda\cdot\frac{a_6(i)}{a_3}}\right)\right) \\
&= \max\left(0, \Lambda\left(1 - \frac{1}{R_0(i)}\right)\right),
\end{aligned} \tag{S27}$$

which is used in the main manuscript and where $R_0(i)$ denotes the reproduction number of strain $i$.

#### Infection probability after wild-type exposure (time-varying drug concentrations, mutational dynamics)

Below, we describe the mathematical approach to compute infection probability after wild-type exposure, with eventual *de novo* drug resistance selection, as shown in Fig. 6 (main manuscript) and Fig. S4.

To compute the infection probability conditioned on a pharmacokinetic trajectory of LEN, we adapt the approach in [13] for computing the probability of extinction and extend it to a multi-strain system comprising wild-type (WT) and mutant strains (taking eqs. (S9)-(S11) as a starting point). The infection probability can be computed as $P_{inf}(Y_0|S_{\text{LEN}}) = 1 - \prod_i P_E(Y_0 = \hat{V}(i)|S_{\text{LEN}})^{V(i)} \cdot P_E(Y_0 = \hat{T}_1(i)|S_{\text{LEN}})^{T_1(i)} \cdot P_E(Y_0 = \hat{T}_2(i)|S_{\text{LEN}})^{T_2(i)}$ where $i$ refers to a mutant strain and $P_E(Y_0 = \hat{V}(i)|\cdot), P_E(Y_0 = \hat{T}_1(i)|\cdot)$ and $P_E(Y_0 = \hat{T}_2(i)|\cdot)$ denote the probability of extinction when exposure to a single free virus $\hat{V}$, early infected cell $\hat{T}_1$ and late infected cell $\hat{T}_2$ of mutant type $i$ occurred and the exponents $V(i), T_1(i)$ and $T_2(i)$ denote the actual size of exposure (assuming statistical independence). Since we consider the infection after exposure to wild-type virus, the equation above reduces to $P_{inf}(Y_0|S_{\text{LEN}}) = 1 - P_E(Y_0 = \hat{V}(\text{wt})|S_{\text{LEN}})^{V(\text{wt})}$.

If only a single variant was present, the extinction probability for this dynamical system can be computed using the following set of ODEs:

$$\frac{d}{dt}P_E\big(Y_t = \hat{V}\big) = (a_1 + a_4)\, P_E\big(Y_t = \hat{V}\big) - a_4\, P_E\big(Y_t = \hat{T}_1\big) - a_1 \tag{S28}$$

$$\frac{d}{dt}P_E\big(Y_t = \hat{T}_1\big) = (a_2 + a_5)\, P_E\big(Y_t = \hat{T}_1\big) - a_5\, P_E\big(Y_t = \hat{T}_2\big) - a_2 \tag{S29}$$

$$\frac{d}{dt}P_E\big(Y_t = \hat{T}_2\big) = (a_3 + a_6(t,i))\, P_E\big(Y_t = \hat{T}_2\big) - a_6(t,i)\, P_E\big(Y_t = \hat{T}_2\big)\, P_E\big(Y_t = \hat{V}\big) - a_3 \tag{S30}$$

where $P_E(Y_t = \hat{V})$ denotes the probability that a single virion present at time $t$ eventually goes extinct. We subsequently shorten this notation to $P_E(\hat{V})$. The pharmacodynamic effect of LEN is included; for brevity, we do not write $S_{\text{LEN}}$ explicitly, but consider propensity $a_6(t, i)$ to be a function of (temporally changing) inhibition by LEN, via the Emax model from eq. (5) (main manuscript). To incorporate mutational dynamics on this system, we couple copies of the above introduced dynamical systems (as shown in Fig 2 (main manuscript) for two strains). Now, the equations for each considered mutant $i$ become:

$$\frac{d}{dt} P_E\big(\hat{V}(i)\big) = (a_1 + a_4)\, P_E\big(\hat{V}(i)\big) - a_4 \left(p_{i\to i}\, P_E\big(\hat{T}_1(i)\big) + p_{i\to j}\, P_E\big(\hat{T}_1(j)\big)\right) - a_1 \tag{S31}$$

$$\frac{d}{dt} P_E\big(\hat{T}_1(i)\big) = (a_2 + a_5)\, P_E\big(\hat{T}_1(i)\big) - a_5\, P_E\big(\hat{T}_2(i)\big) - a_2 \tag{S32}$$

$$\frac{d}{dt} P_E\big(\hat{T}_2(i)\big) = (a_3 + a_6(t, i))\, P_E\big(\hat{T}_2(i)\big) - a_6(t, i)\, P_E\big(\hat{T}_2(i)\big)\, P_E\big(\hat{V}(i)\big) - a_3 \tag{S33}$$

where mutation probabilities $p_{i\to j}$ are defined as above.
While the system above models the probability of extinction (and its complement, the infection), we observed that in cases where mutational escape occurs, that the mutants will always be selected once LEN concentrations are within the mutation selection window. This circumstance is highlighted by the dark shaded areas in Fig. 6 (main manuscript) and in Fig. S4.

#### Mutant selection window (MSW) of once-yearly IM LEN PrEP

Using phenotypic parameters from Supplementary Table S4, we calculated the MSW of once-yearly IM LEN (Study 5, formulation F1) for single, double and triple mutants, as shown in Supplementary Fig. S2 (MSW for twice-yearly SC LEN see *Methods*). Clinically relevant steady-state concentrations for IM LEN once-yearly were 51.40-205.68ng/mL (population-average), with a variability range of 7.7-822.72ng/mL. At average drug levels, Q67H and N74D would be fully suppressed, whereas the lowest concentration (7.7ng/mL) overlaps with their respective MSWs ($[0.8, 21]$ and $[1.7, 59]$ ng/mL, respectively). The MSWs of Q67H+T107N ($[1.9, 141.6]$ ng/mL) and Q67H+K70R ($[1.3, 106.8]$ ng/mL) overlap with the average drug levels of once-yearly IM LEN, while M66I+T107A ($[3.6, 537.2]$ ng/mL) completely overlaps with the average steady-state range. The full concentration range, including inter-individual variability, is entirely contained within the MSWs of Q67H+N74D and Q67H+K70R+T107N ($[2.5, 1107.6]$ and $[0.8, 1142.3]$ ng/mL, respectively).

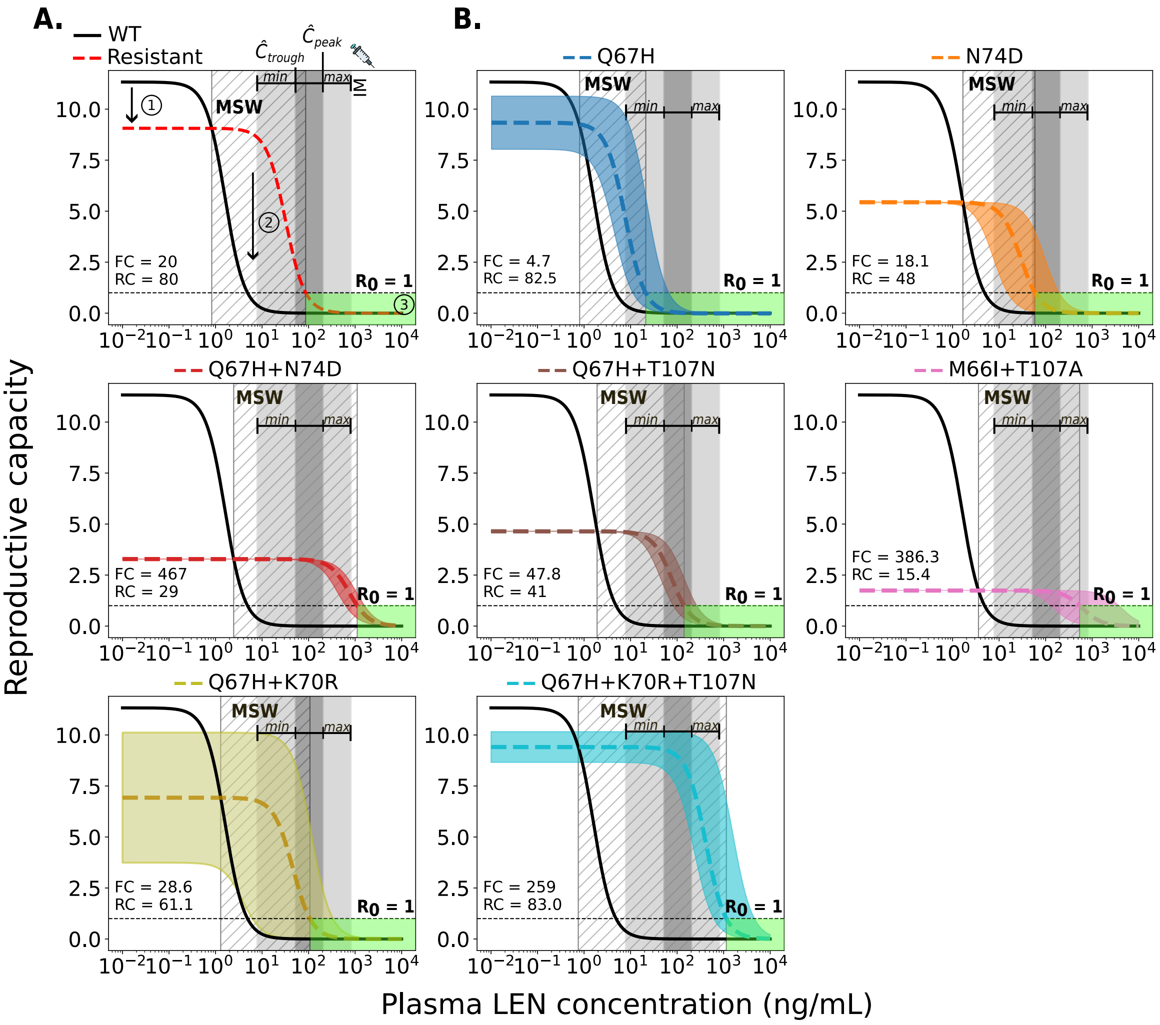


**Figure S2. Relationship between plasma LEN concentrations after once-yearly IM injections and viral reproductive capacity for wild-type and resistant strains.** A. Example showing the impact of a resistant strain on viral fitness (red dashed line: 20% lower replication capacity) and drug susceptibility (solid black line: 20-fold higher $IC_{50}$) compared to wild-type (WT). Key features: (1) reduced fitness of the mutant, (2) the mutant selection window (MSW, diagonal hatching) where the mutant outcompetes WT, and (3) the green area where reproductive capacity is $\leq 1$ (infection cannot occur). B. MSW analysis of two single mutants, four double mutants, and one triple mutant. The horizontal dashed line at $R_0 = 1$ marks the threshold between viral suppression (below 1) and sustained replication (above 1). The dark gray areas indicate the clinically relevant population-average steady-state concentration ranges ($C_{trough}$-to-$C_{peak}$ concentration ranges) achieved by once-yearly IM LEN injections (Study 5, F1). The light gray shaded areas represent ranges of inter-individual variability in drug concentrations informed by observations in the Purpose 2 trial (*min-max*). Each mutant (labeled at the top of each panel) is depicted as a colored dashed line, with uncertainty in fitness and/or fold-change represented by a corresponding shaded region.

#### Prophylactic efficacy of once-yearly IM LEN against WT and mutant HIV strains

As shown in the Supplementary Fig. S3, infection with the WT virus would be completely avoided at the average steady-state LEN concentrations achieved by IM dosing once a year. We investigated how IM-based LEN PrEP (Phase III trial not yet initiated; NCT07047716) may facilitate the transmission of drug-resistant viruses (for details, see Supplementary Table S4). Infection with the single mutant Q67H may occur at the lower variability limit (light gray area; ∼18% median prevention) but would be completely prevented at average steady-state drug levels of IM dosing (dark gray area). Infection with the N74D mutant can also occur, but its median infection probability is reduced by 78-100% on average, with a minimum reduction of at least 10% compared with WT in the absence of LEN. The double mutant Q67H+T107N is not efficiently prevented at IM concentrations (median infection-risk reduction of 14% at the lower variability range; ∼24% at $C_{trough}$) but would be fully prevented at peak steady-state drug levels ($C_{peak}$). The double mutant Q67H+K70R is partially suppressed, with ≥26% median risk reduction at trough levels and complete prevention at peak IM levels. For M66I+T107A, the median risk reduction remains moderate (53–59%) and ranges from 53–100% across the IM variability interval. In contrast, infections associated with Q67H+N74D and Q67H+K70R+T107N are not fully prevented across the IM concentration range, with median risk reductions of 24-65% and 2-51%.

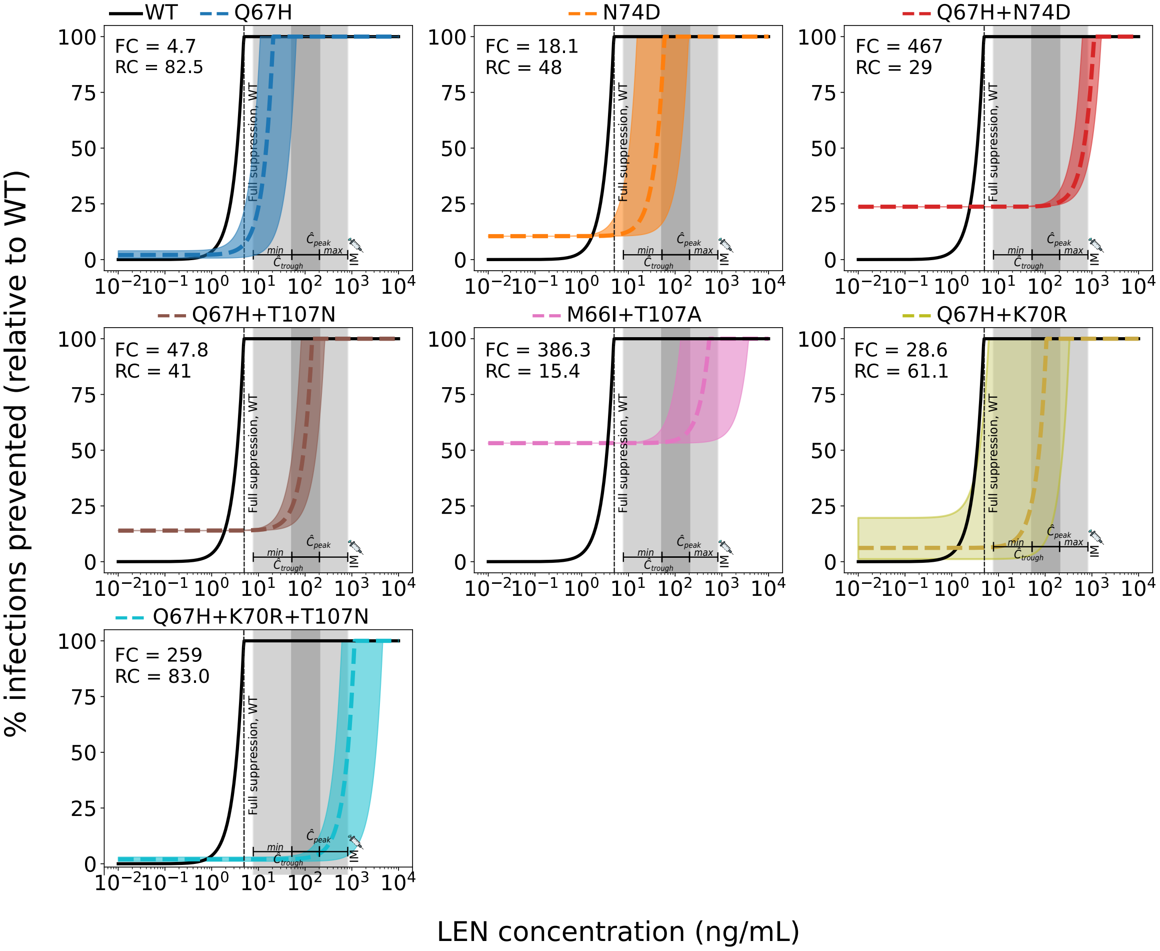


**Figure S3. Reduction in HIV infection risk by IM LEN for wild-type and mutant strains.** Reduction in HIV wild-type (WT) infection risk (black solid lines) and mutant viruses (colored non-solid lines) for single, double and triple mutants. Variability in mutant-specific fitness and/or fold-change values is shown as colored shaded areas corresponding to each mutation, as labeled at the top of each panel. Infection risk reduction (y-axis) with a particular variant and drug concentrations is computed relative to the infection risk with the WT in the absence of drug. The dark-gray area indicates the steady-state concentration range in an 'average individual' ($C_{trough}$-to-$C_{peak}$ concentrations) achieved with once-yearly IM LEN injections. The light-gray area represent ranges of inter-individual variability in drug concentrations informed by observations in the Purpose 2 trial (*min*-*max*). Complete suppression of WT virus is achieved at a LEN concentration of 5ng/mL (dashed vertical line).

#### De novo emergence of resistant strains after LEN IM injection

To compare the relative likelihood of de novo mutation emergence, we considered a time window starting at day 730 and ending at day 1700 after the last LEN IM injection. Approximately 570 days after the last injection, infection with wild-type virus becomes possible and may lead to the de novo emergence of Q67H, N74D, the double mutants Q67H+N74D, Q67H+T107N, M66I+T107A, Q67H+K70R and the triple mutant Q67H+K70R+T107N (see Supplementary Fig. S4). The estimated time windows for de novo selection were approximately 302, 249, 202, 233, 155, 280 and 345 days for Q67H, N74D, Q67H+N74D, Q67H+T107N, M66I+T107A, Q67H+K70R and Q67H+K70R+T107N, respectively. The triple mutation Q67H+K70R+T107N appears to be the most likely mutation to be selected, followed by Q67H, Q67H+K70R, N74D, Q67H+T107N, Q67H+N74D and M66I+T107A. Identical results were obtained for twice-yearly SC LEN injections (see *Results*).

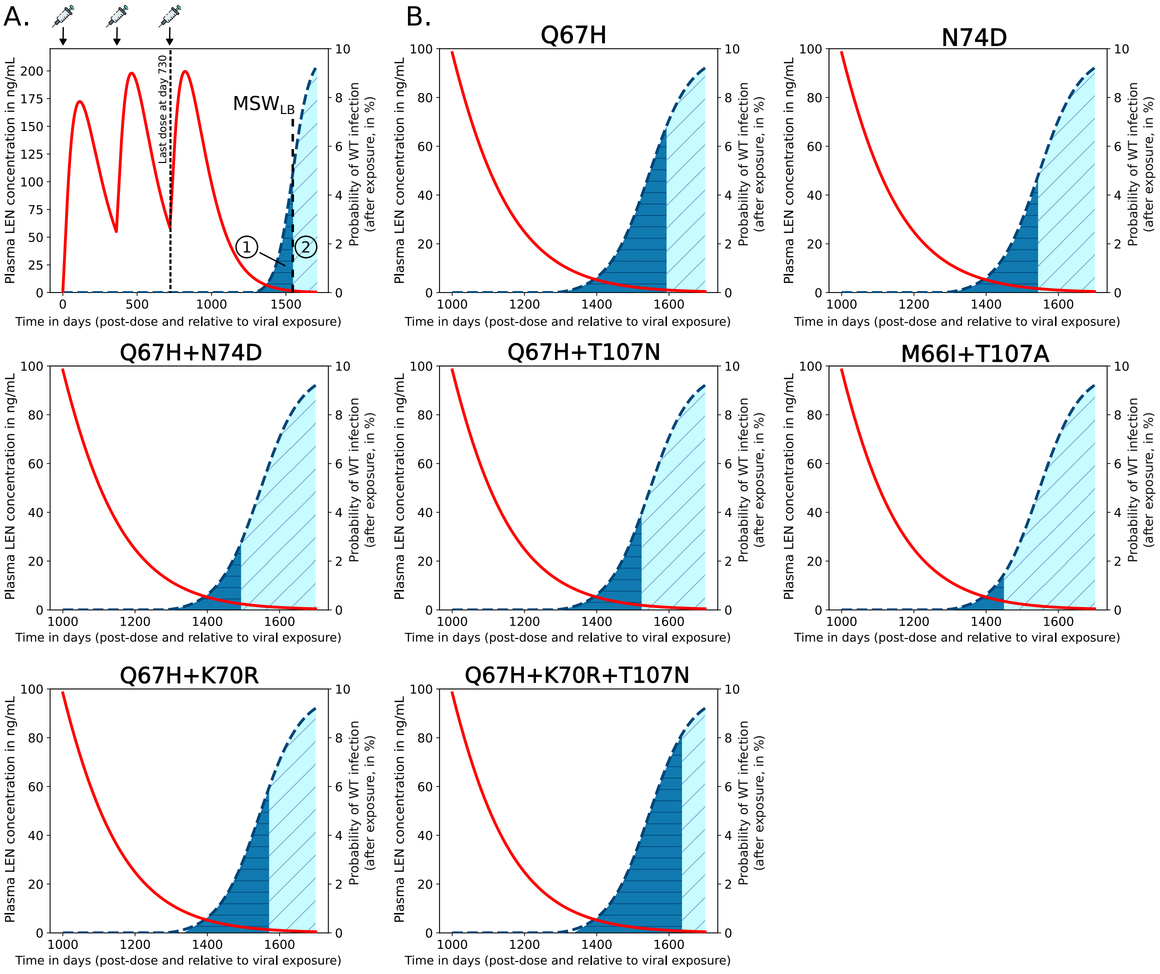


**Figure S4. Quantification of de novo drug resistance emergence risk in scenarios where once-yearly LEN IM doses are missed or when LEN-PrEP is stopped.** Predicted average LEN plasma concentrations (red curve; left y-axis) after the first IM LEN injection and the probabilities of infection if exposure with WT virus occurred at the indicated time after the last LEN injection (blue dashed line; right y-axis). A. Example of once-yearly LEN IM dosing scenario, indicated at the top of the figure. If exposure with WT virus occurs after stopping LEN, two outcomes are possible: (i) infection with WT virus and de novo emergence of a resistant mutant (dark blue area), or (ii) infection with WT virus and selection of WT. The vertical line indicates the lower concentration threshold of the mutant selection window ($MSW_{LB}$, compare Supplementary Fig. S2), i.e. at drug concentrations below this line WT will be selected. B. Simulation results for LEN-associated single, double and triple mutants.

4. Rebecca Begley, Justin Lutz, Martin Rhee, Hadas Dvory-Sobol, Anna Chiu, Steve K West, Jessica Corpus, John Ling, and Polina German. Lenacapavir sustained delivery formulation supports 6-month dosing interval. *AIDS*, 2020.
